# Lack of univariate, clinically-relevant biomarkers of autism in resting state EEG: a study of 776 participants

**DOI:** 10.1101/2023.05.21.23290300

**Authors:** Adam J. O Dede, Wenyi Xiao, Nemanja Vaci, Michael X Cohen, Elizabeth Milne

**Affiliations:** Department of Psychology, University of Sheffield, Sheffield, UK; Department of Medical and Social Sciences, Northwestern University, Chicago, USA; Sincxpress Education SRL

## Abstract

Mental health conditions are difficult to diagnose, requiring expert clinicians and subjective judgements. There has been interest in finding quantitative biomarkers using resting state electroencephalogram (EEG) data. Here, we focus on resting state EEG biomarkers of autism. Although many previous reports have pointed to differences between autistic and neurotypical participants, results have often failed to replicate and sample sizes have typically been small. Taking a big-data, open-science approach, we combined data from 5 studies to create a large sample of autistic and neurotypical individuals (n=776) and used high-power computing to extract 942 variables from each participant’s data. Using a systematic, preregistered analysis pipeline, we failed to identify even a single EEG-based variable that could serve as a practically useful biomarker of autism clinical diagnosis. Our results highlight that a biomarker for autism drawn from EEG data is an elusive construct that may not exist.

## INTRODUCTION

Autism spectrum disorder is a complex, heterogeneous condition diagnosed on the basis of behavioural symptoms.^1^ Although it is widely acknowledged that autism has a neural origin, the precise aetiology of the condition remains elusive. Within the last three decades, many investigations have compared metrics derived from brain imaging techniques of individuals with autism and neurotypical (NT) individuals. A major goal of these investigations has been to identify biomarkers that may prove useful in autism diagnosis and intervention targeting, to shed light on the pathophysiological mechanisms that underlie autism symptoms and to provide grounds for animal back-translation.^2–4^ The search for a biomarker that could be used to diagnose or to screen for autism is particularly attractive as current diagnostic methods are time-consuming, often involving multiple clinical teams, and are fundamentally subjective even when standardised procedures are used.^3^ For example, in one study involving 1814 autism participants, expert clinicians exhibited heterogeneity in their clinical diagnoses despite having the same standardised measures available to them.^5^

While various techniques have been used to obtain data that supports this endeavour, EEG is one of the more commonly used methods because it is cheap, suitable for a range of participants, including infants, is readily available and capable of providing quantitative measurements.^4^ The search for EEG biomarkers of autism is therefore an important research area that has attracted substantial grant funding.

Resting state EEG is of particular interest because it provides direct measurement of neural dynamics without requiring participants to engage in any particular cognitive task. Even small amounts of data (e.g. a five minute recording or less) enable computation of a range of variables, including the power and peak frequency of EEG oscillations, inter-site phase clustering (ISPC), multiscale entropy (MSE), phase-amplitude coupling (PAC), and the slope of the aperiodic power spectrum. Each of these variables reflects dynamics that are contingent on neural architecture and brain biochemistry and therefore may be sensitive to differences between autistic and neurotypical individuals.

Despite substantial work in this area, and the publication of studies that have investigated the above variables in EEG, to date, no clear biomarker has been identified. Instead, the field is characterised by pockets of tantalising observations but no clear conclusions that have been robustly replicated. For example, an early modern study of EEG power spectral density found that autistic children exhibited relatively lower alpha power (8-12 Hz) and relatively higher delta power (1.5-3.5 Hz) than neurotypical children (both groups 6-12 years old), and that autistic children were not significantly different from neurotypical toddlers (2 years old).^6^ This finding, combined with the general fact that resting power spectra shift towards higher frequencies with maturation,^7^ supported the mechanistic explanation that autism may be a form of delayed maturation. However, although some authors have reported replication of increased delta and lowered alpha,^8^ others have failed to replicate the result, and yet others have found evidence for the opposite pattern.^9, 10^ Other frequency ranges have yielded similarly mixed results. The number of studies that have reported an increase in gamma-band power in autism is roughly equal to the number that have reported decreases.^11^

Conflicting results have been reported for other EEG variables as well. For example, autistic participants have presented with both flatter and steeper slopes of the aperiodic power spectrum (1/f trend slope) when compared with neurotypical participants.^12^ Peak alpha frequency has been reported to be higher in children with autism than in neurotypical children,^13^ but also lower in autistic children than in neurotypical children.^14^ Similarly, some autistic participants have exhibited greater PAC between theta phase and gamma power than neurotypical individuals,^15^ other autistic participants have exhibited increased PAC between alpha and gamma oscillations,^13^ but yet other autistic participants have exhibited specific regional decreases as well as increases in PAC between alpha and gamma oscillations.^16^ Autistic individuals generally exhibit decreases in low frequency connectivity,^17^ but some studies have reported increases,^18^ and complex patterns of changes in network connectivity topology have also been reported.^19, 20^ MSE has been reported to be lower in autistic participants than neurotypical participants,^21, 22^ but increased MSE has also been reported.^23, 24^ Perhaps unsurprisingly, although the field appears to be confident that differences do exist in the temporal dynamics of the EEG in autistic individuals compared with neurotypical individuals, no diagnostically relevant biomarkers have emerged.^25^

There are at least five issues that have prevented the field from coalescing around any candidate biomarkers. First, studies typically have small sample sizes, which can lead to estimates of effect sizes that are not reflective of the general population. Second, bias to report significant findings coupled with non-representative effects can lead to systematic bias in the literature.^26^ Third, differences between autistic and neurotypical individuals may develop with age,^13, 14, 22, 27^ and prior studies have often been too small to consider age rigorously. Fourth, in any EEG study, there are always many variables that may be extracted for analysis, yet most investigations report only one or a small handful of variables. As a result, the myriad measures of brain dynamics available in the EEG are treated independently, and it is difficult to evaluate which variables are of most importance. Unless a study and its analysis pipeline have been preregistered, it is difficult to know whether the variable reported in any study is the only one that was examined or if null results were simply put in the “file drawer.” Fifth, a biomarker must reliably improve diagnosis accuracy in a clinical setting. This is a much higher bar than a significant group-comparison difference, yet simple group-comparisons remain the norm in the field.

Although the field lacks consensus about which variable to focus on as a potential EEG biomarker of autism and faces great challenges, there is faith that a useful biomarker of autism does exist in the resting-state EEG. For example, a recent paper stated, “significant evidence supports the notion that developmental differences…measured via resting-state EEG…may be…a suitable biomarker of…ASD diagnosis.” (page 2)^28^

Here, we sought to address these issues and to carry out a systematic investigation of the many EEG variables that have been reported as showing a group difference between autistic and neurotypical individuals by combining data from 5 previously collected datasets to create a large sample of 776 participants across a wide developmental age range (3 to 250 months [21 years]), reporting effect sizes regardless of significance across a large range of EEG variables (942 variables in total), and evaluating diagnosis classification performance of candidate biomarkers using signal detection theory methods that have long been standard for the evaluation of medical tests,^29^ but have rarely been applied to the study of autism.^30^ Data were split into three age groups to simplify analysis and increase sensitivity to differences that may be apparent only during limited developmental periods. Using this approach, we replicated many classic developmental findings in resting-state EEG, and autistic and neurotypical participants exhibited significant differences on several metrics of brain dynamics. However, the metrics that were most different between groups were not ones that would have been easily predicted a priori, and none of these metrics performed well in signal detection theory evaluation. We conclude that there are no clinically relevant biomarkers of autism in the eyes open resting-state EEG within the set of variables that we explored.

## RESULTS

### Qualitative assessment of the data

Figure 2 displays group averaged power spectra and multi-scale entropy within each age group at electrodes Fz, Pz, and Oz. It also displays group average topographical maps of alpha power within each age group and for each diagnosis. Visual inspection of these plots indicates that indices appear as expected, providing reassurance regarding the integrity of the data and the appropriateness of the analysis approach on data that were obtained from several sources. In addition, there were no obvious differences between groups, further motivating our more general approach to searching for group differences.

**Figure 1.**
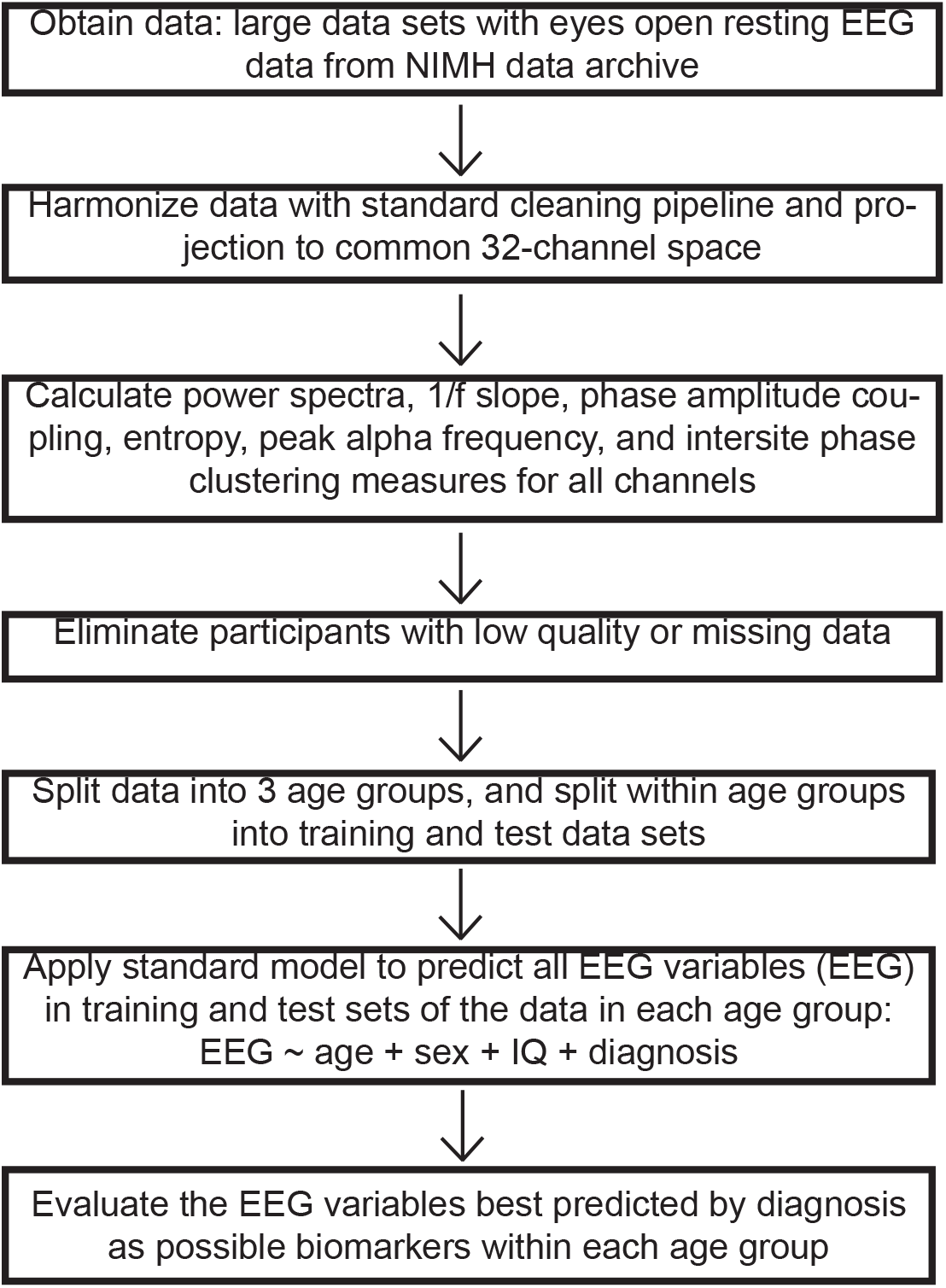
Outline of data search and processing pipeline.

**Figure 2.**
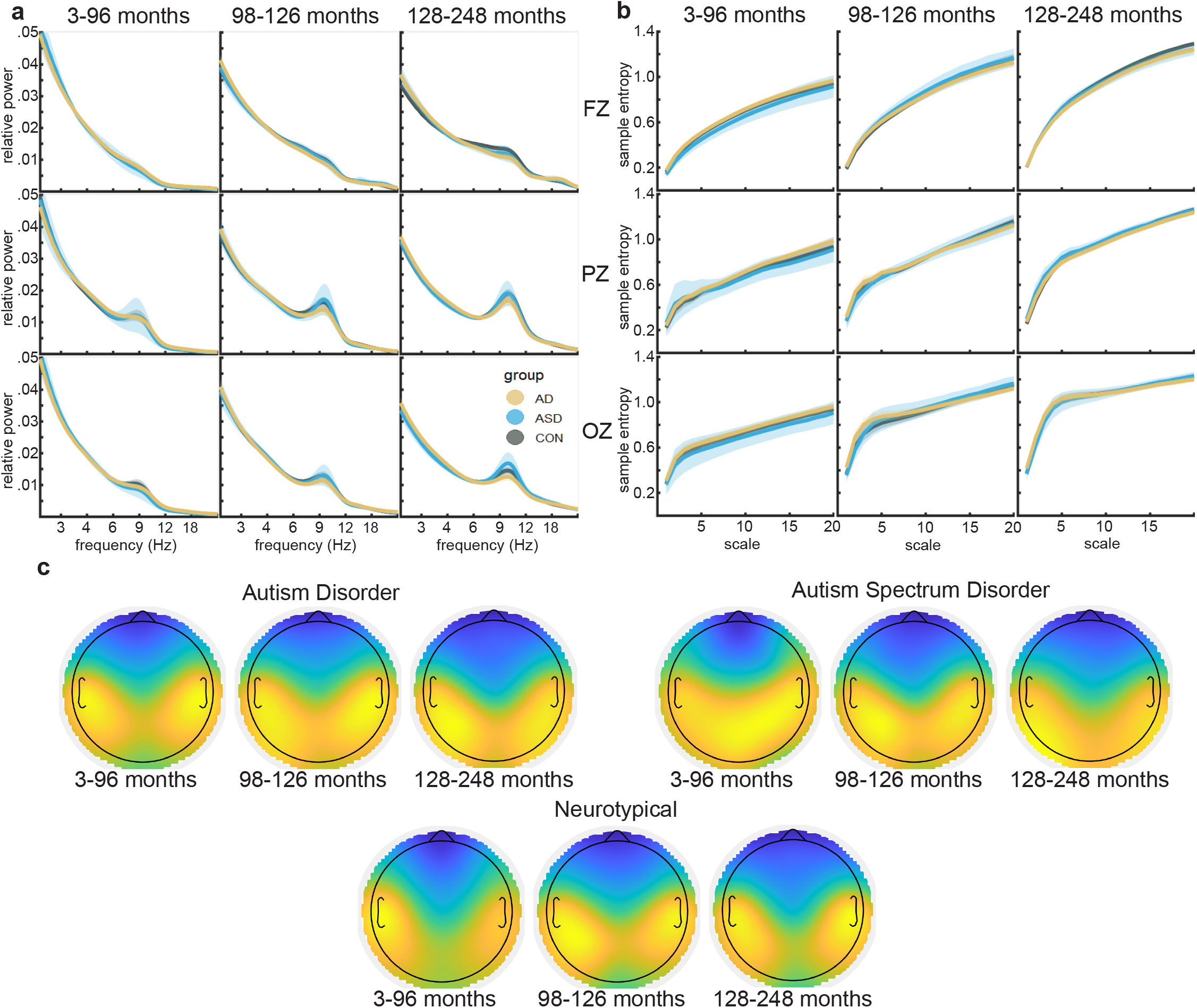
Example group means of selected dependent variables. **a.** Group mean power spectra are displayed in units of relative power. The rows correspond to different electrodes. The columns correspond to age groups. Electrodes and age groups are as labeled. Diagnosis group is indicated in color. Shaded regions indicate the 95% confidence interval. **b.** Group mean multi-scale entropy (MSE) values are displayed. Rows and columns are organized similarly to panel **a**. **c.** Topographical maps of spectral power in the alpha band (7.5 - 14 Hz) are displayed for the three age and diagnosis groups. Each plot is scaled independently to display maximum contrast.

### Quantitative search for optimal candidate biomarkers

For formal analysis, targeted summary values were derived from the calculated EEG measures. Specifically, for each EEG measure, regional averages and difference scores were calculated within the delta, theta, alpha, beta, low gamma, and high gamma frequency bands for 18 different combinations of electrodes. Each combination was chosen to capture a different general region of the scalp. In total, this process yielded 942 dependent variables of interest from each participant. Data were split into three evenly sized age groups. Within age group, data were further split into evenly sized train and test sets and multiple linear regression models were fit for each of the dependent variables in each age group and in the train and test set independently. All model fits used the same four independent variables: age, sex, IQ, and autism diagnosis group. The autism diagnosis group variable had three levels: control (CON), milder autism spectrum disorder (ASD), and more severe autism disorder (AD) (see methods). Extreme outliers were removed prior to fitting the model for each EEG variable (see methods for description of outlier detection procedure). During model fitting, 66.4% of all model fits involved zero outlier rejections. The maximum number of outliers was 27 observations. All instances where there were more than 5 outliers involved fitting either PAC or power variables. There was no difference in the number of outliers as a function of diagnostic group. The mean number of neurotypical outliers removed from each analysis was 0.7 participants, and the corresponding value for autistic outliers was 0.5 participants. Eliminating a small number of outliers should have biassed results towards finding larger effect sizes by limiting sample variability.

The goal of the analysis was to assess which dependent variables could best be predicted by which independent variables. As such, the key output statistic for this analysis was the 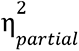 associated with each independent variable in each model fit. This measure of effect size reflects the proportion of the variance unaccounted for by other independent variables that can be accounted for by the independent variable under consideration. That is, when all other variables are already in the model, what proportion of the remaining variance can a particular predictor account for? In addition, a stability index was calculated for all effect sizes. The stability index values reflected the normalised difference between the effect sizes observed for the train set and the corresponding effect sizes observed for the test set.

Critically, since the goal was to identify which dependent variables might serve best as potential biomarkers of autism, it was of interest to identify those independent variables for which diagnosis had an effect size that was both high and stable. To this end, effect sizes were only considered meaningful if they were above .05, and effect sizes were only considered stable if they had stability values of .8 or higher.

Figure 3a displays histograms of the effect sizes associated with different predictors in different age groups. Visual inspection of Figure 3a indicates that age strongly predicted a large number of dependent variables in the youngest and oldest age groups with effect sizes ranging up to .6. Far fewer dependent variables were strongly predicted in the middle age group. The power of sex as a predictor increased across age groups with larger effect sizes being observed for the older two age groups than for the youngest group. By contrast, neither IQ nor diagnosis exhibited strong prediction of EEG dynamics as reflected by a paucity of effect sizes above .05. Panel b of Figure 3 shows the stability of effect size measures. Stability is displayed for effect sizes that were greater than .05 in the training data set only. Visual inspection of these plots reveals that stability values for age and sex were generally high with distributions for both independent variables grouped towards the maximum value of 1.0. By contrast, stability values for IQ and diagnosis were generally low with distributions for both independent variables grouped towards the minimum value of 0.0.

**Figure 3.**
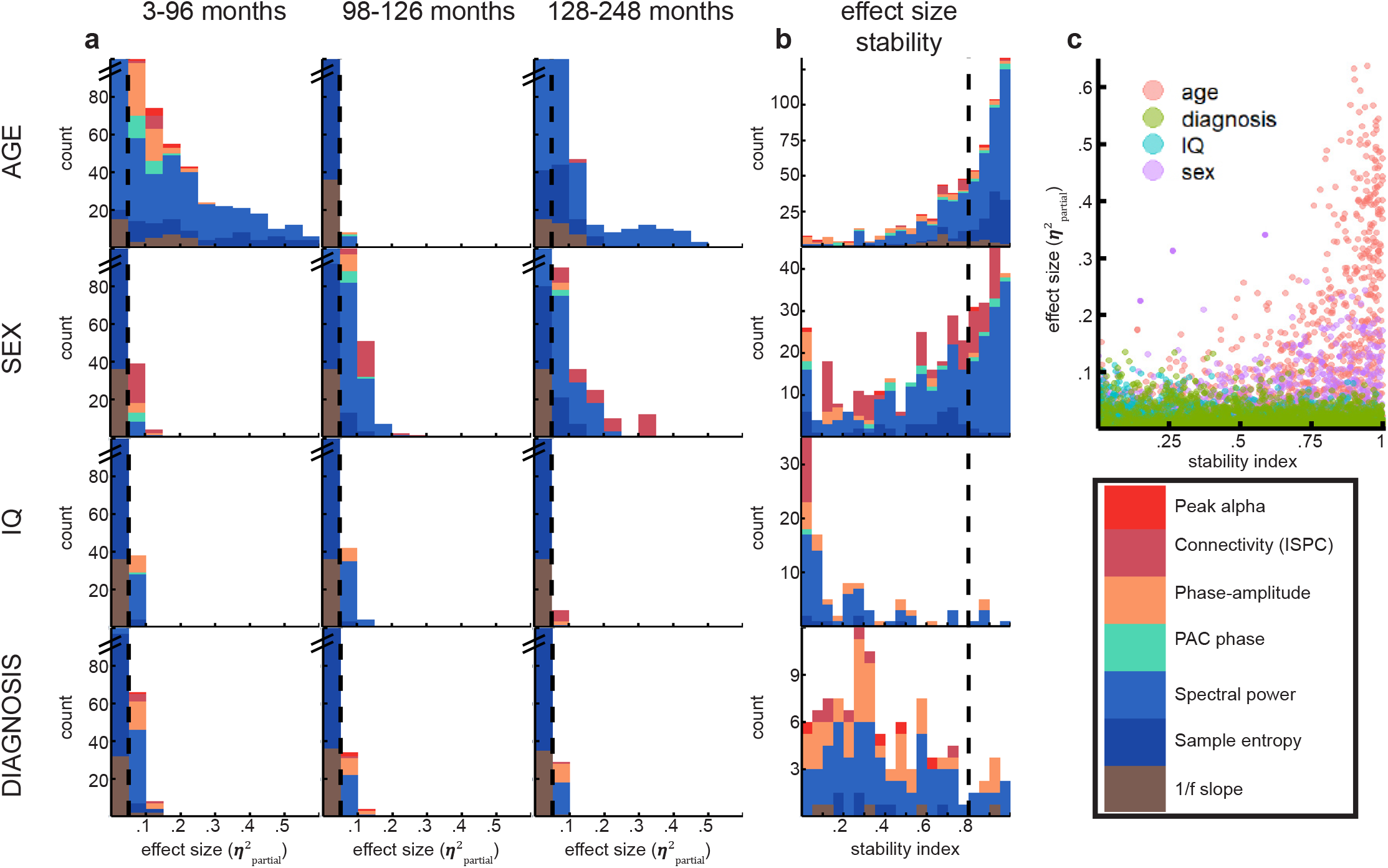
Age and sex were good predictors of resting state EEG, but IQ and diagnosis were not. **a.** Each histogram displays the effect sizes observed for the independent variable indicated at left within the age group indicated at the column head. Colors indicate the type of dependent variable being predicted (see key). Note that all histograms have truncated y axes in order to focus on larger effect sizes. Dashed vertical lines at .05 indicate the threshold used to separate meaningful effect sizes from noise. **b.** Each histogram displays the stability index for effect sizes greater than .05. The stability index was calculated by comparing corresponding effect sizes in independently analyzed training and test datasets. The stability index ranges from 0 to 1 where larger values indicate higher stability. Colors indicate the type of dependent variable being predicted (see key). All age groups are combined. The vertical dashed lines at .8 indicate the threshold used to separate stable from unstable effect sizes. See table 2 for counts of the number of dependent variables that were well predicted by each independent variable in each age group. Well-predicted was defined as a prediction that exceeded both the effect size and stability thresholds. In general, far more dependent variables were well-predicted by age and sex than by IQ or diagnosis. **c.** The scatter plot displays the relationtionship between effect size and stability. For age and sex there is a positive correlation, but there is no positive correlation for diagnosis or IQ.

Quantitatively, Table 3 displays the number of dependent variables that were predicted with an effect size greater than .05 and a stability index greater than .8 for each independent variable within each age group. Supplemental Table 1 provides the full list of EEG variables that were well-predicted by diagnosis as well as a breakdown of the beta weights associated with AD and ASD groups relative to CON. Age was a strong and stable predictor of the most measures of EEG dynamics, followed by sex, diagnosis, and IQ. The number of well-predicted EEG variables was not distributed evenly across age and independent variable (*χ*^2^(6)=227, p<<.0001). To assess individual predictors further, the relationship between stability and effect size was evaluated for each predictor (Figure 3c). For age and sex, effect size was positively correlated with stability (r values > .24; p values <<.0001), indicating that larger effect sizes in the training set were more likely to replicate. By contrast, this correlation was missing for IQ (p=.53). It was reversed for diagnosis (r=-.06; p=.003). In other words, there was a small but significant relationship such that larger diagnosis prediction effects were more likely to be unstable and fail to replicate.

**Table 1:** Demographic and data quality statistics for all participants

| group | data set | IQ | IQ metric | age | n female | n total | orig channels | final channels | orig epochs | final epochs |
| --- | --- | --- | --- | --- | --- | --- | --- | --- | --- | --- |
| AD | biomarkCon | 98 (18) | DAS GCA | 106 (20) | 39 | 168 | 124 (0) | 109 (15) | 107 (121) | 78 (11) |
| ASD | biomarkCon | 108 (13) | DAS GCA | 107 (21) | 7 | 23 | 124 (0) | 117 (8) | 91 (0) | 83 (8) |
| CON | biomarkCon | 116 (12) | DAS GCA | 104 (20) | 35 | 106 | 124 (0) | 117 (8) | 117 (151) | 85 (8) |
| AD | biomarkDev | 71 (24) | MSEL | 39 (38) | 10 | 68 | 125 (0) | 89 (17) | 52 (16) | 40 (13) |
| ASD | biomarkDev | 106 (19) | MSEL | 11 (8) | 4 | 10 | 125 (0) | 90 (12) | 50 (18) | 36 (4) |
| CON | biomarkDev | 104 (17) | MSEL | 9 (4) | 22 | 50 | 125 (0) | 92 (13) | 53 (14) | 41 (12) |
| AD | femaleASD | 99 (20) | WTAR | 146 (32) | 41 | 112 | 125 (0) | 125 (1) | 65 (27) | 65 (27) |
| ASD | femaleASD | 103 (21) | WTAR | 155 (38) | 25 | 40 | 125 (0) | 125 (1) | 67 (29) | 67 (29) |
| CON | femaleASD | 113 (15) | WTAR | 156 (35) | 92 | 178 | 125 (0) | 125 (1) | 80 (23) | 80 (23) |
| AD | socBrain | 99 (10) | DAS GCA | 233 (9) | 0 | 7 | 62 (0) | 62 (0) | 154 (4) | 124 (34) |
| ASD | socBrain | 97 (12) | DAS GCA | 233 (13) | 0 | 3 | 62 (0) | 51 (9) | 152 (3) | 101 (44) |
| CON | socBrain | 97 (0) | DAS GCA | 232 (6) | 1 | 2 | 62 (0) | 62 (0) | 154 (6) | 132 (23) |
| AD | bpSZ |  | WTAR |  | 0 | 0 |  |  |  |  |
| ASD | bpSZ |  | WTAR |  | 0 | 0 |  |  |  |  |
| CON | bpSZ | 104 (9) | WTAR | 216 (21) | 4 | 9 | 62 (0) | 59 (3) | 100 (0) | 79 (10) |

DAS GCA = Differential Ability Scales General Conceptual Ability
MSEL = Mullen Scales of Early Learning
WTAR = Weschler Test of Adult Reading
biomarkCon = The Autism Biomarkers Consortium for Clinical Trials
biomarkDev = Biomarkers of Developmental Trajectories and Treatment in ASD
bpSZ = Bipolar & Schizophrenia Consortium for Parsing Intermediate Phenotypes
femaleASD = Multimodal Developmental Neurogenetics of Females with ASD
socBrain = The Social Brain in Schizophrenia and Autism Spectrum Disorders
Numeric values indicate mean and (standard deviation).

**Table 2:** ages are reported in months. Parenthetical values indicate the number of female participants in each group

| group | age min | age max | n AD | n ASD | n CON |
| --- | --- | --- | --- | --- | --- |
| Youngest | 3 | 96 | 123 (26) | 19 (7) | 93 (36) |
| Middle | 98 | 126 | 115 (31) | 21 (9) | 84 (42) |
| Oldest | 128 | 248 | 108 (29) | 35 (19) | 161 (73) |

**Table 3:**
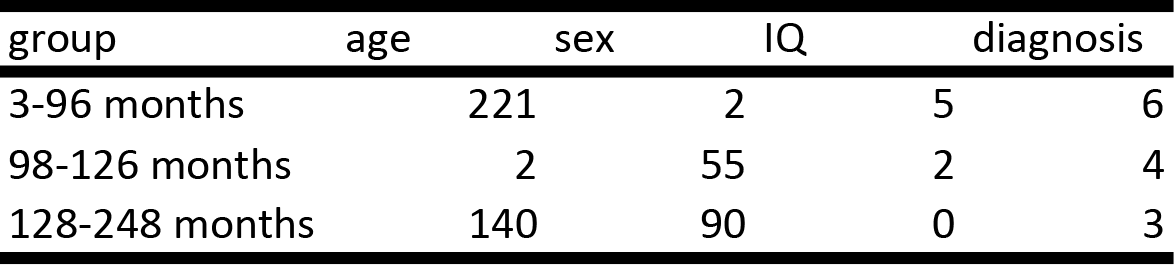
Performance of different predictors in each age group Number of independent variables predicted with η^2^_p_ > .05 and stability > .80

| group | age | sex | IQ | diagnosis |
| --- | --- | --- | --- | --- |
| 3-96 months | 221 | 2 | 5 | 6 |
| 98-126 months | 2 | 55 | 2 | 4 |
| 128-248 months | 140 | 90 | 0 | 3 |

**Table 4:** Signal detection theory measures of biomarker classification performance.

| Age Group | AUC | Accuracy | Sensitivity | Specificity | IG (.0148) | IG (.187) | FPI (.0148) | FPI (.187) |
| --- | --- | --- | --- | --- | --- | --- | --- | --- |
| 3-96 mon | 0.55 | 0.57 | 0.53 | 0.60 | 0.00 | 0.04 | 52.33 | 3.33 |
| 98-126 mo | 0.62 | 0.62 | 0.49 | 0.76 | 0.01 | 0.13 | 32.73 | 2.14 |
| 128-248 m | 0.65 | 0.63 | 0.45 | 0.81 | 0.02 | 0.17 | 26.52 | 1.79 |

AUC = area under the receiver operating characteristic curve
IG (.0148) = information gain when base rate is .0148
FPI (.0148) = number of false positives for each true positive when base rate is .0148
IG (.187) and FPI (.187) = corresponding values for a base rate of .187

### Signal detection theory assessment of candidate biomarkers

The three variables found to be best predicted by diagnosis (one for each age group, variables displayed in Supplemental Figure 2g-i) were evaluated for their effectiveness as biomarkers of autism. These variables will be referred to as “biomarkers” throughout this analysis. For this analysis, AD and ASD groups were combined. ROC curves were plotted (Figure 4a), and the area under the curve (AUC), accuracy, sensitivity, and specificity were all calculated in each age group (Table 4). In general, across groups, classification accuracy was below 65%.

**Figure 4.**
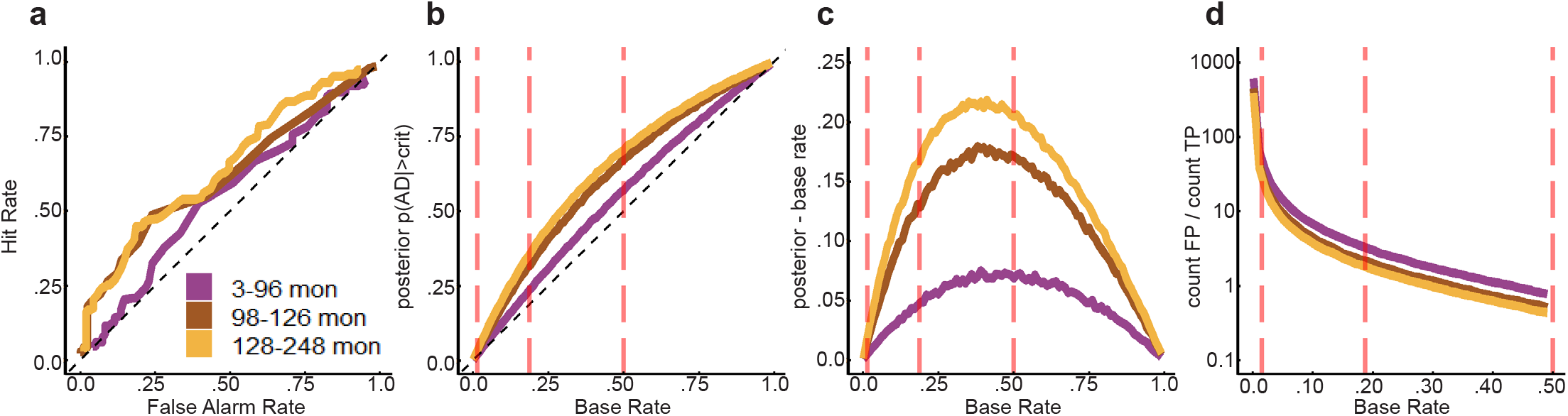
Data driven selection of best possible candidate biomarkers yielded poor biomarker performance. All data from both training and test sets were pooled for this analysis. The variables best predicted by diagnosis (variables displayed in panels **g-i** of Figure 4) were used as biomarkers to detect autism independently in each age group. Signal detection theory-based classification was used to evaluate biomarker performance. Colors indicate different age groups (see key). **a.** The line plot displays the receiver operating characteristic (ROC) curves associated with detection of autism (AD/ASD) within each age group. The hit rate is displayed on the y-axis and the false alarm rate is on the x-axis. A hit was defined as a correct identification of a participant with autism. A false alarm was defined as falsely labeling a control participant as one with autism. ROCs near the dashed diagonal corespond with chance performance. For panels **b-d,** the x-axis displays the base rate of individuals with autism in the population. Data were simulated by sampling with replacement from the observed data to obtain different target base rates. Vertical red dashed lines in panels **b-d** indicate base rates of autism prevalence of theoretical interest (see main text). **b.** The line plot displays classification performance across various different simulated base rates. The y-axis displays the posterior probability that a participant has autism given that they were labeled as having autism according to the biomarker classifier. The dashed diagonal indicates chance performance. **c.** The line plot displays the information gained by application of the biomarker classifier. The y-axis indicates the difference between the posterior probability and the base rate. That is, this plot displays the difference between the solid lines and the dashed diagonal in panel **b**. **d**. The line plot displays the false positivity index associated with different base rates. The y-axis displays the ratio of false positives to false negatives, termed the false positivity index. Conceptually, this index is the number of false positives one can expect to observe for each true positive when applying a biomarker classifier.

In our sample, 55.6% of participants were in the AD or ASD groups (see Table 2). For a biomarker that could be used in a clinical situation, this base rate may not always reflect the base rate of the population undergoing diagnostic screening. Thus, we simulated results from 100,000 participants. Simulated participants were drawn randomly with replacement from the empirical distributions of observed EEG measurements. Simulations were run with varying proportions of participants drawn from the distribution of AD/ASD participants vs. the distribution of CON participants. The varying proportions reflected different simulated base rates for autism in the population. For each simulation, we calculated the posterior probability of a participant having been drawn from the empirical autism distribution given that they were categorised as autistic. The prior for this calculation was the base rate of autistic participants simulated in the data. In other words, chance performance of the biomarker would be a posterior value that precisely equals the base rate. Figure 4b displays the values of the posterior, as a function of the simulated base rate for each age group. These curves indicated that the candidate biomarkers could increase the posterior probability above the level anticipated by the base rate alone. To visualise this more specifically, information gain was also plotted (Figure 4c). Information gain represents the difference between the posterior and the base rate prior. In both Figure 4b and 4c, it is clear that information gain varies as a function of base rate. Base rates near 0 or 1.0 lead to situations in which the biomarker is relatively less useful. We also calculated the false positive index (FPi; Figure 4d). The FPi is the ratio of false positive cases to true positive cases. Conceptually, this measure assesses how many false positives one can expect for every true positive. This value also varies heavily with base rate. When the base rate is low, the number of false positives for each true positive becomes extremely high. In the real world, a biomarker that yields 10s or even 100s of false positives for each true identification is unlikely to be useful.

To quantify classification performance, we repeated the simulated analysis described above 100 times at each of three base rates: .0148, .187, and .5. These base rates were chosen because they reflect an estimate for the population prevalence of autism,^31^ the prevalence of autism in children who have a sibling diagnosed with autism,^32^ and the typical proportion of participants with autism in scientific studies of autism. These base rates are indicated by vertical dashed lines in Figures 4b-d and the information gain and FPi for these base rates are presented in Table 4. Through repeating these simulations, it was possible to assess stability around estimates of AUC, information gain, and FPi. AUC, information gain, and FPi were all extremely stable across simulations. ANOVAs were run for AUC, information gain, and FPi. Each ANOVA used age group, base rate, and the interaction between these as factors. For AUC, there was a main effect of age group (F(2, 891)=31,619.7; p<<.0001), but no interaction or effect of base rate (ps>.05). In other words, while AUC varied as a function of age group, it was not sensitive to base rate. By contrast, for information gain, both main effects and the interaction were significant (Fs(2/4, 891)>34,000; p<<.0001). For FPi, both main effects and the interaction were significant (Fs(2/4, 891)>13,000; p<<.0001). These results validate the conclusions derived from visual inspection of Figure 4. It is important to note that AUC is similar and highly correlated to categorization accuracy measures employed in many studies, and it is not sensitive to base rate. By contrast, information gain and FPi have never been reported with respect to autism diagnosis; they are extremely sensitive to base rate, and they have more relevance to clinical performance of the biomarkers than AUC or accuracy.

## DISCUSSION

The search for reliable resting-state EEG biomarkers that can identify autistic individuals is a growing area of research. A limitation of much of the existing literature in this area is that studies are generally small, i.e. include fewer than 100 participants, and report only a small number of variables. Subsequently, the risk of both type 1 and type 2 statistical errors is high and no reliable biomarkers for autism have yet been identified. Here, we capitalised on the foresight and generosity of researchers and funding agencies that have collated and shared existing datasets, by obtaining and pooling resting-state EEG datasets from autistic and matched samples of neurotypical individuals. We analysed data from 776 individuals, 421 of whom were diagnosed with AD/ASD. We took an exploratory approach and, after applying a standardised pipeline to harmonise and clean the data, used established methods to extract multiple variables that have previously been suggested as potential candidates for autism biomarkers, or to show group differences between autistic and neurotypical samples. We compared each variable between autistic and neurotypical samples to identify which EEG variables were most different between groups and may have utility as potential biomarkers.

We found little evidence that autism could be reliably predicted by any of the EEG variables we examined. The majority of effect sizes were small and/or inconsistent across training and test data sets, and there was a small negative correlation between effect size and stability of effect (Figure 3). Only 13 out of 2826 variables (942 considered independently in 3 age groups) passed our threshold of being noteworthy by virtue of having a medium effect size (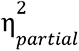 >.05), and a stability index of > .8. No unique variable passed this threshold in each of the three age groups (table S1). Further, as can be seen in supplemental Figure 2g – i and in our signal detection theory evaluation of candidate biomarkers, even for the variables that were most different between diagnosis groups, the AD, ASD and CON participants exhibited substantial overlap.

Despite finding little evidence of differences between the autistic and neurotypical samples, our results replicated previous findings with respect to the range of values obtained for key variables and the topographical distribution of alpha power across the scalp (see Figure 2), indicating that combining distinct datasets acquired from different labs is a valid approach. More specifically, we replicated age-related increase in MSE^33^ and age-related decrease in theta and delta power^7, 34, 35^ (see Supplemental Figure 2a - c), supporting both the integrity of the data and the validity of our analysis approach. Furthermore, associations between age and EEG variables yielded more large effect-sizes (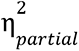 > .1) in the youngest age group, when brain development is known to be rapid, and the sizes of these effects were correlated with their stability (Figure 3), providing further reassurance of the data and the methods. We also found a number of large and stable effect sizes associated with sex as a predictor, particularly in the middle and oldest groups. As sex differences in EEG is not the focus of the current paper, these were not explored further, but these findings suggest that this is worthy of further investigation. We found that IQ was largely unrelated to the EEG variables (see table 3). It is important to note however that the vast majority of participants had IQ within the normal range. Furthermore, since the different datasets included here used different instruments to measure IQ (Table 1), the ability to detect clear associations between particular EEG variables and IQ may have been reduced.

A critique of the search for autism biomarkers, as highlighted by Waterhouse,^36^ is the claim that there is limited biological and construct validity of the diagnosis of autism. The results reported here speak to this position given the striking lack of reliable differences in EEG data between the autistic and neurotypical individuals. Cognition, behaviour and perception are underpinned by large-scale neural dynamics which are reflected by the EEG variables measured here. Therefore, if the alterations in the expression of cognition, behaviour and perception that define autism arise from differences in neural networks supporting these functions it would be likely that this would be apparent via clear differences in at least some of the variables measured. It may be that an autism diagnosis, as it is currently defined, cannot be identified with sufficient consistency to yield a consistent neurological profile across individuals with the diagnosis.

A related concern in the search for biomarkers for autism is the impact of heterogeneity of the condition.^37, 38^ As highlighted by Lombardo,^39^ if autism is, in fact, a broad description for what may be a constellation of unique genetic and neurological conditions, then particular biomarkers may reflect specific subsets of autistic individuals, but not reliably discriminate all individuals with an autism diagnosis from a neurotypical sample. Indeed, inspection of table S1 reveals that the beta weights associated with AD and ASD were sometimes quite different, suggesting a variable relationship between diagnosis and EEG dynamics as a function of symptom severity. Further, recent work has demonstrated that grouping participants by differences in genetic copy number variations (CNVs) can more effectively predict fMRI connectivity than grouping the same participants by mental health diagnosis, based on behavioural symptoms and experiences.^40^ This work implies that clearer neural differences will be seen when groups are based on a known CNV than on a diagnosis where potential genetic variance is unknown. The genetics of autism are complicated and largely unclear, with many candidate genes implicated.^41^ Genetic information was not available for the datasets analysed here, therefore it was not possible to refine analyses based on specific CNV abnormalities. Future research will likely benefit from searching for biomarkers within homogeneous subgroups of autistic individuals.^42^ Nevertheless, if the goal is to identify a candidate EEG biomarker for autism as it is currently defined and diagnosed, which was the goal of the present research and of myriad published and on-going studies, our results cast doubt on the likelihood of achieving this goal and the value in pursuing this endeavour further.

Our results are based on resting-state EEG data. We therefore cannot rule out the possibility that clearer EEG differences between autistic and neurotypical individuals may manifest during task engagement. Neither can we rule out the possibility that there are further variables that can be extracted from the resting state EEG signal that may more successfully differentiate the autistic from the neurotypical sample. Nevertheless, the approach we took to analysis was comprehensive and included indices that represent fundamental features of neural dynamics, therefore it would be somewhat surprising for a novel measure to demonstrate meaningful univariate group differences.

There are a growing number of studies that have reported high levels of classification accuracy using multivariate, machine learning approaches to classify EEG data obtained from autistic and neurotypical samples. However, these studies typically include a much smaller number of participants than is reported here,^43^ and there is no clear consensus on the appropriate classification algorithms or discriminative features of the EEG that lead to such high levels of classification accuracy. What’s more, to our knowledge, no multivariate algorithm for autism diagnosis has ever been applied to a dataset outside the one on which it was developed. Although studies frequently use leave-one-out or k-fold cross validation, it is known that when analyses are repeated multiple times during development, modelling approaches become adaptive, leading to overfitting.^44, 45^ This risk is particularly high for approaches based on support vector machines when the number of features is substantially higher than the number of participants, a situation that is common to machine learning approaches to autism diagnostic classification.^46^ The importance of truly independent samples for training and testing of diagnostic classification is also highlighted by the present finding that the largest effect sizes associated with differences between diagnostic groups were also the most unreliable between the training and test sets. This raises the possibility that high classification accuracies presented in previous papers may be based on the idiosyncrasies of particular datasets. A strength of the present study is that we preregistered our core statistical model, and we did not alter the model in search of greater significance. We also randomly split our data into training and test sets one time on a particular calendar day, further limiting our ability to adaptively fit the data. Future machine learning studies will likely benefit from the larger datasets that are now being made publically available which allow for more rigorous statistical approaches. In addition, future machine learning studies will also benefit from adding signal detection theory analysis to the evaluation of their algorithms.

Finally, our conclusion that there are no univariate, clinically-relevant biomarkers of autism in the resting-state EEG is based on our application of signal detection theory, which is not typically used in studies of autism. We found differences between autistic and neurotypical participants (supplemental figures 2g-i) as large as many of the differences reported throughout the autism literature, and although our signal detection theory analysis (Figure 4) demonstrates that effects of this magnitude cannot serve as useful biomarkers, this has not stopped them from being interpreted as biomarkers. For example, the N170 event related potential has been submitted to the FDA’s Biomarker Qualification Program.^47^ However, setting aside points that have been made previously about how the N170 varies across development, has uncertain correlation to behaviour, and is highly variable between individuals,^48^ the N170 is likely to be a poor biomarker because the effect size of the group difference between autistic and neurotypical participants is in a range where the present analysis indicates that the information gain will be too low and the false positive index too high to make a meaningful clinical difference in real world diagnosis. A recent paper from the same group to propose the N170 as a biomarker reported an 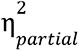 value of .084 for a diagnosis X task condition interaction.^49^ Setting aside the difficulty of using an interaction effect as a diagnostic test, we have demonstrated here that when signal detection theory methods, taking into account naturalistic base rates, are applied to effects of this magnitude, classification performance is poor.

In summary, our results highlight that a univariate biomarker for autism, in so far as can be drawn from eyes-open resting state EEG data, is an elusive construct that may not exist. Future research should focus on analysis approaches that control for heterogeneity between autistic individuals as it may be more fruitful to attempt development of biomarkers for genetically, physiologically, or phenotypically identified subgroups than for autism in general.

## METHODS

This project was pre-registered,^50^ however alterations were made to the analysis plan in order to better summarise the data and test the hypothetical assumptions. Differences and their justifications are noted below. The overall analysis pipeline as finally implemented is shown in Figure 1.

Due to the size of the dataset analysed here, analysis depended on efficient use of high power computing (HPC) resources provided by the University of Sheffield. Please see the supplement for details regarding structuring analysis code and formatting data for compatibility with HPC analysis.

### Data

No new data were collected for this project. We combined data from 5 separately collected datasets (Table 1). All raw data were obtained through the National Institute of Mental Health (NIMH) data archive (NDA), and can be accessed there by any interested researchers.^51^ With the exception of the bpSZ dataset, all autistic participants were evaluated using the ADOS module appropriate to their age and language ability. Neurotypical participants were assessed using either the ADOS or the judgement of qualified clinicians in the data collection teams. Where two modules of the ADOS were collected for the same participant, we used only the score resulting from the module designed for the lower language ability. For this analysis, only data from control participants in the bpSZ dataset were considered. These participants were deemed not to have any major psychological disorders by the clinicians involved in the bpSZ dataset’s collection. For participants whose EEG had been collected multiple times, only their first EEG dataset was considered. Only participants with eyes open resting state data were considered. For participants with both eyes open and eyes closed data, only eyes open data were considered. All participants were sorted into one of three groups based on ADOS score and defined using the terminology provided by the ADOS: an Autism Spectrum Disorder (ASD) group, a more severe Autism Disorder (AD) group, and a control (CON) group. The motivation for splitting the ASD group into an ASD and an AD group was in recognition of the heterogeneity of the condition; ADOS score was the only information available across all participants to support any kind of sub-typing. This sorting was based on standard cut offs as specified in the ADOS use manual.^52^ Across datasets, after applying the above inclusion/exclusion criteria, there were 1040 participants. In the preregistration, we had planned to analyse data from participants across the entire lifespan. However, there were relatively few observations of individuals with autism above the age of 250 months (380 autistic individuals less than or equal to 250 months old; 12 autistic individuals greater than 250 months old). Thus, analysis was limited to participants less than or equal to 250 months old, leaving 808 participants. Also different from the pre-registered plan was the division of the data into age groups. Specifically, for analysis, participants were split into 3 age groups, and the cut offs between these age groups were determined such that one third of the AD and ASD participants were included in each group. This was done because visual inspection of several EEG metrics as a function of age revealed non-linearity that would have required changing our pre-registered modelling plan. Age cut offs were determined after rejection of noisy data sets (see below).

Our pre-registration did not anticipate missing IQ data, but for 9 participants, IQ data were missing. For these participants, a value equal to their dataset and diagnosis group mean was substituted.

### Preprocessing

Preprocessing and cleaning followed standard methods which are reported in detail in our preregistration^50^ and in the supplemental methods.

### Extraction of key EEG variables

Power spectra, 1/f trend slope, peak alpha frequency, phase-amplitude coupling (PAC), multi-scale entropy (MSE), and intersite phase clustering (ISPC) were all calculated using standard methods. For details please see supplemental methods and our preregistration.

### Channel groupings for comparisons

All calculations generated values for every data channel (except ISPC, which generated values for pairs of channels). However, signals at adjacent electrodes are correlated. In addition, utilising all calculated values as dependent variables would be intractable. Thus, the preregistration plan was augmented with the following electrode groupings and frequency bands for statistical analyses.

The following 13 regional groupings were used: right frontal (FP2, AF4, F4, F8), left frontal (FP1, AF3, F3, F7), right centroparietal (FC2, FC6, C4, CP2, CP6), left centroparietal (FC1, FC5, C3, CP1, CP5), right occipito parietal (P4, P8, PO4, O2), left occipito parietal (P3, P7, PO3, O1), frontal (FP1, FP2, AF3, AF4, F4, FZ, F3), occipital (PO4, PO3, O2, OZ, O1), central (FZ, CZ, PZ, OZ), left lateral (F7, FC5, T7, CP5, P7), right lateral (F8, FC6, T8, CP6, P8), right hemisphere (FP2, AF4, F4, F8, FC6, FC2, T8, C4, CP6, CP2, P8, P4, PO4, O2), left hemisphere (FP1, AF3, F3, F7, FC5, FC1, T7, C3, CP5, CP1, P7, P3, PO3, O1).

Comparisons of particular asymmetries have also featured in the resting state autism literature.^53 54^ Thus, we designed 5 asymmetry sensitive comparisons. In these comparisons, the second channel in each pair is being subtracted from the first, and the difference is divided by the sum of the two values (except for comparisons involving ISPC where the calculated values already represented pairs of electrodes). The mean of each set of subtractions was calculated. Specifically, we used the following sets: interhemispheric asymmetry: FP1-FP2, F3-F4, F7-F8, C3-C4, T7-T8, P3-P4, P7-P8, O1-O2; intrahemispheric asymmetry in the rostrocaudal direction in the left hemisphere: O1-P3, P3-C3, P7-T7, C3-F3, T7-F7, CP1-FC1; intrahemispheric asymmetry in the rostrocaudal direction in the right hemisphere: O2-P4, P4-C4, P8-T8, C4-F4, T8-F8, CP2-FC2; intrahemispheric asymmetry in the mediolateral direction in the left hemisphere: P7-P3, CP5-CP1, T7-C3, FC5-FC1, F7-F3; intrahemispheric asymmetry in the mediolateral direction in the right hemisphere: P8-P4, CP6-CP2, T8-C4, FC6-FC2, F8-F4. Note the importance of maintaining consistency in caudal, rostral and lateral, medial ordering across all pairs in these comparisons.

For power spectrum comparisons, spectra from each channel were averaged in 6 frequency bands: δ (2-4 Hz), θ (4-8 Hz), α (8-14 Hz), β (14-30 Hz), γlow (30-50 Hz), and γhigh (50-80 Hz). These averaged band power values were then used to calculate all regional and asymmetry comparisons. This process was repeated for raw, log-transformed, and relative power values. There were 324 (18 comparisons X 6 frequencies X 3 measures) dependent measures for power spectra.

For 1/f trend slope the regional and asymmetry comparisons were calculated for slope values derived from both log-transformed and relative power values. There were 36 (18 comparisons X 2 measures) dependent measures for 1/f trend slope.

For peak alpha frequency the regional and asymmetry comparisons were calculated for peak alpha frequencies derived from both log-transformed and relative power values. There were 36 (18 comparisons X 2 measures) dependent measures for peak alpha frequency.

For PAC, low frequencies were divided into δ (2-4 Hz), θ (4-8 Hz), α (8-14 Hz), β (14-20 Hz), and high frequencies were divided into β (20-32 Hz; note when β was compared to β, 24 Hz was used as the low cut off for the higher frequency), γlow (32-52 Hz), and γhigh (52-100 Hz). Thus, there were 12 pairs of frequency bands, and PAC was averaged within the range of each frequency band for every channel. Average PAC values were calculated for all regional and asymmetry comparisons. This process was repeated for PAC values z-scored with respect to their shuffle-generated null distributions, and for phase preference values. There were 432 (18 comparisons X 12 frequency pairs X 2 measures) dependent measures for PAC.

For MSE, scale was divided into four ranges: all (1-20), fine (1-7), medium (8-13), coarse (14-20). MSE within each channel was averaged across these scale ranges. These averaged MSE values were used to calculate all regional and asymmetry comparisons. There were 72 (18 comparisons X 4 measures) dependent measures for MSE.

Finally, for ISPC, frequency was divided into the same 6 bands as above for the power spectrum. Within each band, the average ISPC was calculated for the electrode pairs specified in the 5 asymmetry comparisons (note: regional comparisons were not possible for ISPC as it requires pairs of electrodes). In addition, the mean long distance connectivity was calculated by taking the mean ISPC across all channel pairs whose inter-electrode euclidean distance was greater than the median inter-electrode euclidean distance. The mean short-distance connectivity was calculated similarly using pairs whose inter-electrode distance was less than the median. There were 42 (7 comparisons X 6 frequency bands) dependent measures for ISPC.

Thus, in total there were 942 (324 power + 36 1/f slope + 36 peak alpha + 432 PAC + 72 MSE + 42 ISPC) dependent measures.

### Statistical evaluation of group differences for every dependent measure

The analysis of the data followed the preregistration plan,^50^ where all measures were regressed onto the age, sex, IQ and autism diagnosis variables.

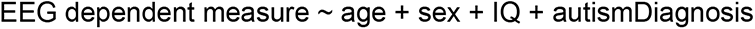

Data were divided into three age groups such that there were an equal number of AD and ASD participants in each group. Specifically, age groups were: 0-8.08 years; 8.16 - 10.58 years, and 10.66 - 20.83 years. After dividing participants into age groups, participants were split into a training and a test set. This split was done such that within each age group, the two sets had an equal total n, the same proportion of AD, ASD, and CON participants and the same proportion of males and females. We used 50% of the data to train the models and and 50% of the data to test and compare them, which is contrary to the proposed 70% - 30% splits in the preregistration document. The change in percentage split of the data allowed us to calculate not only effect sizes but also measures of effect size stability across the two halves of the data. Random assignment to train and test set was done once on the 29th of November, 2022, and the necessary data for all analyses were saved as separate .csv files for each set. Because randomisation was performed only once, it was not possible to bias the results by repeating randomisation and recalculating output statistics after each repetition.

We performed 942 statistical tests within each age group and on each half of the data, resulting in a total of 5652 tests. The coefficients were estimated in the R statistical environment (R core team, 2021) using linear regression (lm builtin function). For each test, 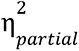 associated with each independent variable (age, sex, IQ, and diagnosis) was calculated to measure effect size. In all cases, type III sums of squares was used to calculate ηpartial2. Using type III sums of squares means that all effect size values reflect the effect of a predictor assuming that all other predictors were added into the model before it. With such a large number of tests, it was guaranteed that many would yield significant results. We were interested in the distribution and stability of effect sizes associated with our four predictors across different dependent measures rather than with significance on any individual test.

Finally, outliers were removed prior to model fitting. This had to be done for each test individually since participants could exhibit outlier values for some EEG dependent variables but not for others. For some dependent variables, outliers were found to be so extreme that a small handful of values could drag the mean far above or below all other observations. In these cases, simply rejecting all observations more than some number of standard deviations from the mean would not work because all observations fell far from the mean. To get around this, the following two step procedure was adopted for each dependent variable. First, the mean and standard deviation of the middle 80% of the observed values were used to z-score all values. In this way, extreme outliers could not contribute to the standardisation. Second, all values with z-scores within 5 standard deviations were used to recalculate the mean and standard deviation for a final z-distribution. The data were then standardised according to this second set of mean and standard deviation values, and data points with z scores of greater than 5 in magnitude were deemed outliers and not included in the analysis.

### Effect size and stability of the effect

To assess the stability of the observed effects, a stability index was calculated for each effect size:

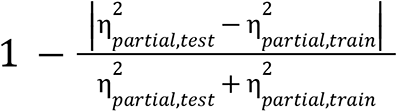

An independent variable was deemed a good predictor of a particular EEG dependent variable if it had a stability index above .8 and an 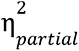 above .05.

We counted the number of EEG dependent measures that could be well-predicted by each independent measure within each age group and used a ^2^ test to assess whether there was a difference in the strength of prediction of the EEG across independent variables and age groups. For each predictor, the correlation between effect size in the train set and the stability index was calculated. Good predictors should exhibit a positive correlation such that larger effect sizes are also more stable.

### Evaluating diagnostic accuracy of EEG measures as biomarkers

Our goal was to evaluate whether or not the EEG variables that differed significantly between the autistic and control participants could serve as useful biomarkers when evaluated using signal detection theory (SDT). To do this, for each age group, out of the set of reliable effect sizes (stability indices > .8) the dependent measure that was best predicted by diagnosis was selected for evaluation as a possible biomarker. A good biomarker needs to advance patient diagnosis in the real world, and an SDT-based analysis provides tools to go beyond simply evaluating the significance of a group difference.

Each candidate biomarker was submitted to receiver operating characteristic (ROC) analysis ^55, 56^ (see Table 3 for further descriptions of all SDT measures). Empirical ROCs were plotted (Figure 6a) by calculating hit and false alarm rates for 100 linearly spaced diagnostic classification criteria values across the range of the candidate biomarker. The area under the ROC (AUC) was calculated analytically. An optimal criterion was chosen by searching for the criterion that yielded the highest average of hit rate and correct rejection rate. This measure was termed accuracy. Specifically, this accuracy measure was calculated as (hit rate + correct rejection rate) / 2. Accuracy was not biassed by base rate. Sensitivity and specificity were evaluated at this criterion. These measures provided metrics of classification performance under idealised conditions.

To emulate real-world conditions, we also evaluated biomarkers with simulations that took into account the base rate of autistic prevalence in the sample.^55, 57^ To do this, we used 100 linearly spaced base rates between .001 and 1. For each base rate, 100,000 participants were drawn randomly with replacement from the observed distributions of autistic and control biomarker values in proportion to the target base rate. Each random sample of data was submitted to SDT analysis using three metrics known to be sensitive to base rate: precision (Figure 6b), information gain (Figure 6c), and false positive index (FPi)(Figure 6d). Precision is the posterior probability of a participant being autistic given that they are categorised as autistic. In this calculation Bayes’ theorem was applied with the relevant base rate serving as the prior. Information gain is precision minus the prior (base rate). Information gain represents the amount of diagnostic information that is gained through the use of the biomarker. FPi is the ratio of false positives to true positives. Effectively, it measures how many false alarms can be expected for each true positive. The lower this value is, the better. In our preregistration, we had not proposed the random sampling based approach. This approach was necessary in order to obtain stable estimates particularly at very low and very high base rate levels.

ASD and AD participants were combined into a single autistic group. Only participants within the age group that each biomarker was detected were used for evaluating each biomarker. For each biomarker, if the autistic group had a lower mean than the control group, then the sign of the biomarker measurement was inverted for all participants to facilitate comparison of detection performance across age groups.

For the youngest age group, the dependent measure associated with the largest effect size appeared to have yielded a large effect size because it was elevated for ASD but not for AD participants. Thus, we evaluated the dependent measure that diagnosis had predicted with the second largest stable effect size. For the other two age groups, the dependent measure associated with the largest effect size had a higher magnitude beta weight for AD than for ASD.

As a further augmentation to the preregistered analysis plan, we assessed the stability of these measures and evaluated formally how they changed with base rate; simulations were repeated 100 times at three specific base rates: .0148, .187, and .5. These values were intended to simulate diagnostic screening in the general population,^31^ diagnostic screening of high-risk children,^32^ and the typical categorization problem encountered in scientific studies with artificially matched group sizes, respectively. For AUC, information gain, and FPi, change as a function of age group and base rate was evaluated using 3 (base rate) X 3 (age group) ANOVAs.

## Data Availability

All EEG variables extracted from the raw data are available at the individual participant level at https://github.com/adede1988/SheffieldAutismBiomarkers.git. All raw data are available on the National Insitute of Mental Health Data Archive website in repository 10.15154/1528473.

## Data Availability Statement

All EEG variables extracted from the raw data are available at the individual participant level at https://github.com/adede1988/SheffieldAutismBiomarkers.git. All raw data are available on the NDA.^51^

## Code Availability Statement

Analysis was performed in Matlab and R. Custom scripts referenced throughout the methods are available at https://github.com/adede1988/SheffieldAutismBiomarkers.git.

## Supplemental methods and results for

Matlab functions referenced throughout this supplement can be found at https://github.com/adede1988/SheffieldAutismBiomarkers.git.

## Data Preprocessing

All data were obtained in a raw state with the exception of the female ASD data set, which was provided on the NDA having already been high pass filtered at .1 Hz, low pass filtered at 100 Hz, and notch filtered at 60 Hz. In addition, channels recorded from electrodes with impedances over 200 kOhm had already been deleted and data were split into 2.048 second epochs by the original data collection team.

For all participants, data were segmented into 2 second epochs (female ASD set left as 2.048 s epochs). Each epoch was forward and backward reflected before being high pass filtered at .5 Hz (matlab function: highpass), low pass filtered at 200 Hz (matlab function: lowpass), and notch filtered at 60 or 50 Hz (depending on country of data collection) to eliminate line noise (matlab builtin functions: iirnotch and filtfilt).

The difference between the maximum and minimum voltage in a moving 80 ms window was calculated for each channel and epoch. Epochs where at least one 100 μV deflection was detected were flagged as potentially noisy. A channel was deemed bad if it crossed the 100 μV threshold in 50% of trials. Bad channels were removed. A trial was deemed bad if 25% of the remaining channels exhibited threshold crossings. Bad trials were removed. These parameters were modelled on the preprocessing steps used in the female ASD data set, which was provided precleaned. In deviation from our pre-registration, we added two data rejection criteria to reject entire participants whose data was deemed too noisy. First, participants whose original data as it came to us included fewer than 20 channels were removed from further analysis. This criterion eliminated one participant. Second, participants for whom 50% or more of channels were rejected were removed from further analysis. This criterion eliminated 31 participants. Thus, the final dataset used for all further analyses included 776 participants. For these participants, the mean and standard deviation of the numbers of trials and channels per participant before (original) and after (final) cleaning are displayed in Table 1. The final numbers of participants included in all groups are displayed in Table 2.

Finally, all data were re-referenced to an average reference and interpolated to a standard 32-channel montage. Interpolating all data to a standard montage facilitated comparison between data collected using variable numbers of electrodes.

For more detail about data import and cleaning steps, see functions readEEGdat.m, removeNoiseChansVolt.m, and convertCoordinates.m.

## Computation of key EEG variables

### Power spectra

Power spectra were calculated independently in each epoch. The epoch was forward and backward mirrored to avoid edge artifacts, and wavelet convolution was applied for 100 logarithmically spaced frequencies between 2 and 80 Hz ^1^. The resulting filtered complex time series was converted into a power time series, the mirrored copies discarded, and its mean was taken across the epoch. After repeating this procedure for all epochs, the mean was taken across epochs. This yielded a 32 (electrodes) X 100 (frequencies) matrix of power values. For more detail, see function getPower.m.

### 1/f trend slope

The 1/f trend slope was calculated for each channel independently using the power spectra calculated above. Calculations followed the procedure outlined in ^2^. Calculations were carried out on both the log-transformed power spectra and the relative power spectra. Power values for frequencies below 7 Hz and between 14 and 24 Hz were used for slope fitting. Values between 7 and 14 Hz were left out to avoid the slope fit being skewed by the alpha peak. A line was fit to the remaining power values as a function of frequency. For more detail, see function getSlopeAlpha.m.

### Peak alpha frequency

Peak alpha frequency was calculated for each channel independently using the power spectra calculated above. Calculations followed the procedure outlined in Dickinson et al. ^3^. Calculations were carried out on both the log-transformed and relative power spectra. Power values for frequencies between 6 and 14 Hz were used for calculation. These power values were detrended by subtracting the fitted 1/f trend line. Next, a gaussian curve was fit to the detrended power spectrum. The mean of this gaussian was taken as the peak alpha frequency. If the model fitting procedure failed to converge or if the mean of the gaussian fell outside the range 6-14 Hz, then this was taken as evidence that the participant did not have a strong alpha peak at that electrode and this participant’s data were not interrogated further with respect to peak alpha frequency at that electrode. Regional means and asymmetry calculations (see below) ignored missing peak alpha frequencies at individual electrodes. 262/776 participants had at least one regional or asymmetry calculation that was not possible due to missing values for all electrodes involved in the calculation. However, no participant was missing more than 18 of 36 regional/asymmetry values. Only 22 participants were missing more than 5 values. In all, 97.7% of all peak alpha dependent measures were successfully fit. For more details, see function getSlopeAlpha.m

### Phase-Amplitude Coupling

Phase-amplitude coupling (PAC) was calculated independently in each epoch and for each channel. Calculations generally followed suggestions in ^4, 5^. To do this, 10 low frequencies were chosen (2:20 Hz with spacing of 2 Hz). These frequencies were associated with linearly spaced standard deviation values of 2 to 3.5. 21 high frequencies were chosen (20:100 Hz with spacing of 4 Hz). These frequencies were associated with linearly spaced standard deviation values of 3.5 to 6. For each combination of low and high frequency, data were filtered using a frequency domain, gaussian-shaped filter convolution method, equivalent to time-domain wavelet convolution ^1^. The low frequency complex time series was converted into a phase time series using the matlab angle function. The high frequency complex time series was converted into an amplitude time series using the matlab abs function. The high frequency amplitude time series was divided into bins on the basis of the low-frequency phase time series. Specifically, the phase angles were split into 18 evenly-sized bins (i.e. 20 degrees per bin). The average high frequency amplitude was calculated for each bin. These mean values were normalized by the sum of the means across all bins.

For comparison, the same PAC calculation was carried out on data where the low frequency phase time series and the high frequency amplitude time series had been randomly temporally shifted relative to one another. In this way, it was possible to calculate the distribution of PAC strength that would be expected by chance. Temporal shifting and recalculation was carried out 200 times to build up a null distribution.

This procedure yielded an epoch X 10 (low frequency) X 21 (high frequency) X 18 (phase bin) X 32 (channel) matrix of observed PAC values and an associated 200 (random temporal shift repeats) X epoch X 10 (low frequency) X 21 (high frequency) X 18 (phase bin) X 32 (channel) matrix of null PAC values. The mean of both matrices was taken across the epoch dimension. Then, the Kullback-Leibler divergence was computed on both matrices across the phase bin dimension relative to a uniform distribution. This yielded matrices of epoch-averaged modulation indices for all combinations of channel, low frequency, and high frequency.

Observed modulation indices were converted into z-scores relative to their corresponding temporally shuffled null distributions.

Phase preference was calculated as the weighted circular mean of the bin phases, weighted by the mean amplitude values observed in each bin phase. Phase preference was calculated after averaging over epochs.

For more detail on how PAC values were calculated see function getPAC.m.

### Multi-Scale Entropy

Multi-Scale sample entropy (MSE) is a method that calculates sample entropy on both the original signal and coarse-grained time series derived from it. The algorithm consists of two steps that are performed independently for each electrode and epoch. First, the original signal is resampled to 1000Hz to ensure consistency across datasets. Then, coarse-grained time series are generated by averaging consecutive data points over time scales that increase with scale factor, resulting in a time series of length exactly divisible by the scale factor.^6^ Second, sample entropy is computed for each coarse-grained time series using a similarity threshold of 30% of the standard deviation of the time series denoted as γ, and a pattern length of *m*.^7^ Before computing sample entropy, all time series are centered and normalized to standard deviation 1 to avoid bias from amplitude variations. The distance between pairwise elements is calculated using the Chebyshev distance metric and the number of thresholded pairwise distances is counted.

and sample entropy equation is:

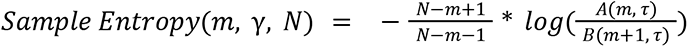

where *m* is the pattern length (fixed at 2), τ is the time scale factor (from 1 to 20 in this study), *N* is the length of the original signal, *A*(*m*, τ)is the number of pairs of vectors with a Chebyshev distance less than or equal to the similarity threshold for scale factor τ and pattern length *m*, and *B*(*m* + 1, τ) is the number of pairs of vectors with a Chebyshev distance less than or equal to the similarity threshold for scale factor τ and pattern length *m* + 1.

This yielded an epochs X 20 (time scales) X 32 (channels) matrix. The mean of both matrices was taken across the epoch dimension. For more detail on how MSE values were calculated see functions getEntropyVals.m, Multi.m, and SampleEntropy.m.

### Inter-Site Phase Clustering

Inter-site Phase clustering (ISPC) was calculated independently for every epoch between all electrode pairs within the same 100 logarithmically spaced frequencies between 2 and 80 Hz that were used for power spectrum calculation. Importantly, data were transformed using the surface Laplacian (custom matlab function laplacian_perrinX.m) prior to ISPC calculation. Calculation generally followed the procedure described in ^1^. Specifically, for each frequency, data were first filtered using the same procedure described above. Next, filtered data were converted into a phase time series. Then, all pairwise channel comparisons were made using the following formula:

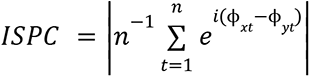

Here, n is the number of time points in the epoch, *i* is 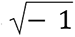, ϕ_*xt*_ is the phase angle of the signal from channel *x* at time *t*, and ϕ_*yt*_ is the corresponding value for channel y. This yielded an epochs X 100 (frequencies) X 32 (channels) X 32 (channels) matrix. Finally, the mean was taken with respect to epoch. For more detail on how ISPC was calculated see function getISPC.m.

## High performance computing methods

Considering the large number of participants whose data contributed to this project and the comprehensive set of variables that were measured for each participant, the use of a high performance computing (HPC) cluster was necessary to complete this analysis within a manageable amount of time. Here, we present key considerations that facilitated the structuring of analysis code and formatting of data in order to use HPC resources effectively. The examples given throughout assume that the reader has downloaded the github repository and is able to view the scripts contained in it (https://github.com/adede1988/SheffieldAutismBiomarkers.git). It is our hope that the functions to compute specific aspects of EEG dynamics will prove useful to other researchers. The scripts and functions written to organize file paths and metadata, and that control loading data and saving outputs are all specific to the present analysis and are intended more as examples than as code that will be useful off the shelf to other researchers.

There are 4 key system considerations when switching from analysis on a local machine to using HPC resources. First, HPC is best suited to applications that require a high number of processors and a low amount of memory per processor. Second, management of HPC inputs and outputs requires a higher level of code and data organization than local analysis. Third, HPC resources are managed by scheduling software that will grant low priority to users who request large amounts of resources frequently. Fourth, because analysis using HPC clusters often involves processing a very large number of inputs, it encourages the user to automate many of the steps that are performed interactively during local analysis. These differences motivate how to structure analyses for optimal use of HPC resources.

### 1. General code structure for interacting with HPC clusters

When doing analysis on one local machine, users are accustomed to opening their preferred analysis software and then executing a series of code lines, function calls, or graphically selected menu options. This process is usually done for each subject and variable to be analyzed separately involving a high degree of customization and interaction throughout the analysis process. Often, the code associated with a project will end up being contained in one or a small handful of long scripts that require user edited file paths and parameter adjustments to work properly. This process can be very time consuming and difficult to keep track of. Analysis on an HPC encourages users to automate interactive steps, which will likely mean structuring code quite differently.

In addition to speeding up analysis, increasing automation when doing HPC analysis also improves memory usage and reproducibility. When analyzing one subject at a time on a local machine, users generally load entire datasets into memory and carry a large number of interim calculated variables through analysis. Although this can facilitate flexibility and interaction, it uses a lot of memory and it can be difficult to reproduce final results that are dependent on interim analysis steps. When analyzing using HPC clusters, it is best practice to load the minimum amount of data into memory at a time and to save interim outputs rather than to keep them loaded in memory.

For the present study, each script or function of analysis code could be categorized into one of four nested layers. Thinking of the code as belonging to these layers helps to understand the overall architecture of the code. Figure S1 displays a schematic representation of the interactions between the four code layers, the raw data, and the saved outputs. Specifically, Figure S1 displays initial multichannel analysis for one of the datasets we analyzed. To best understand this example, the relevant code files are available in the SheffieldAutismBiomarkers github repository. The example scripts are specific to the Autism Biomarkers Consortium for Clinical Trials dataset. As will be explained below, only layers 1 and 2 of the code were dataset specific. Layers 3 and 4 were dataset general. This structure allowed new datasets to be analyzed using the same computation code (layers 3 and 4) even if they required different formatting code (layers 1 and 2).

The defining feature of the code layers was that the code in the top layer was called from the command line on the HPC by the user. Code in each lower layer was called from the immediately higher layer. The top layer consisted of bash scripts, and we termed this layer the job initialization layer. These scripts described to the HPC cluster the computing resources required, the number of jobs to be done, the analysis software (Matlab in the present case) to be used for each analysis job, and the specific initializing commands to be sent to the Matlab command line to begin each job (see exampleBashScript.sh). One of these initializing commands was for Matlab to run one of the scripts belonging to the second layer.

When creating job initialization bash scripts, it is important to consider the system requirements of the particular HPC being used for analysis. The Sheffield Advanced Research Computer (ShARC) used for the present analysis relied on a job input and scheduling program called SLURM (or simple linux utility for resource management). SLURM is a linux operating system scheduling program. It is commonly used on HPC systems. If implementing an HPC analysis on an HPC system that also uses SLURM, then it will be possible for users to use the same command structures shown in our exampleBashScript.sh. However, different HPC systems can use different scheduling and resource management systems, so it is important to check for compatibility as job initialization bash scripts may need to be adapted for the particular HPC that they are intended to run on. In most cases, research computing support staff are highly knowledgeable and helpful in properly formatting these initialization scripts.

The second layer consisted of pipeline wrapper scripts. The pipeline wrappers contained file paths to locate data and code and to specify where outputs should be saved. Depending on the application, they also specified the output format and populated metadata for the output. A key aspect of the wrappers was that they did not open any data files, and their main goals were formatting, job monitoring, and job initiation. They did no computation. The only truly necessary feature of any pipeline wrapper script was that it needed to call a pipeline function, the third code layer. The example script included in the github repository (getBioConsortDat.m) is written for the initial processing of the Autism Biomarkers Consortium for Clinical Trials dataset. It draws on a .csv file called biomarkConDat.csv to obtain metadata about participants. This file is also available in the github repository. The metadata were used to initialize the summary statistics file which contained interim output variables and was passed back and forth throughout the rest of the analysis (see green arrows in Figure S1).

The third layer consisted of pipeline functions. The pipeline loaded the raw data and then submitted the data to a series of computation steps to extract target variables. The pipeline kept output variables organized using the summary statistics file specified in the wrapper. Finally, the pipeline saved the summary statistics file (see setReadInAggregate.m).

A key difference between the pipeline wrapper and the pipeline itself was that the pipeline wrapper always included a directory of all relevant data files for the entire analysis, but it didn’t actually open any data. By contrast, the pipeline dealt only with one data file path, and it did open those data. The main advantage of this separation was that multiple different wrappers could be combined with a single pipeline. This was critical to standardize analysis across different datasets. For example, note that the getBioConsortDat.m script is written for one specific dataset. Similar scripts were written for each dataset taking into account the specific idiosyncrasies of each dataset’s file and data organization. No matter these differences, each wrapper script formatted file paths and metadata into the same format and then made a final call to the same pipeline (setReadInAggregate.m in this example). An additional advantage was that it was easy to change pipeline scripts while keeping the same wrapper(s), which facilitated analysis code development. Basically, the wrapper script answered the questions: which data are to be analyzed, where is the code to analyze them, and where should outputs be saved? The pipeline function then loaded the data, oversaw the analysis, and saved outputs.

Finally, the fourth code layer consisted of analysis computation functions. Each computation (cleaning, intersite phase clustering, power spectra, 1/f slope etc.) was contained in a separate computation function. These computation functions were highly modular. Thus, it was easy to add or subtract computation functions from the analysis pipeline. Packaging computations into separate subfunctions also had the advantage of making the pipeline functions highly human readable. Most of the functions provided in the github repository are computation functions that extract measurements of particular features of the EEG. Each computation function took in data and summary statistics. They all saved summary statistics and processed data in addition to sending output back to the pipeline.

The result of these layers was that the code library for this analysis included many different code files, each with a highly specialized job.

### 2. Development of HPC analysis code

A few key concepts can help dramatically in developing analysis code using the multi-layer architecture used in the present study. Here, we describe four coding conventions that helped in the present project: file path control, debugging, modularity in code organization, and frequent output saving and progress checking. It is important to note that the code included in the github repository is intended primarily as an example that is specific to the present analysis.

Implementing a similar analysis on a different HPC for a different analysis will require significant customization.

#### 2.1 File path control

During development, code was written on a local machine and only deployed on the HPC cluster after completing debugging. Working locally during development facilitated easy interaction with the code and its outputs during development. The major challenge with this workflow is file path control.

Even when it is possible to utilize a network attached storage drive that can be viewed from both the HPC interface and one’s local machine, the file paths used during HPC and local processing will be different. To get around this issue, file paths should be specified dynamically rather than statically. It is likely that in any scenario, it will only be the first part of any file path that will change between HPC and local processing. Thus, it is possible to specify the different part of all file paths as a single variable. For the present study, we used the convention that this variable was named “prefix”. Note the variable named “prefix” specified at the top of getBioConsortDat.m. As a general rule, the prefix used for local processing should always be contained in a comment, and the prefix used for HPC processing should be in uncommented code. This will allow an interactive local user to run the local prefix but will prevent accidental file path incompatibility when moving to HPC analysis. Combining this convention with GitHub to keep code aligned between local machine and HPC cluster allowed for seamless movement between local and HPC analysis. In addition to using a prefix variable to standardize file paths, as a general rule, file paths should only be specified in job initiation and pipeline wrapper scripts. This rule makes file path errors easy to trace and fix.

#### 2.2 Finding errors and debugging

Errors could occur during HPC processing for one of two general reasons. First, as with local processing, errors occurred because of mistakes in the analysis code. Second, errors could occur because of process failure on the HPC cluster.

Taking code errors first, it was important to be able to reproduce errors that occurred during analysis on the cluster on a local machine. It was much easier to examine the causes of errors when they could be reproduced on a local machine. To do this, errors had to be localized to the code layer (section 1) and particular script or function in which they occurred. Although there is no Matlab graphic user interface (GUI) when running analyses using an HPC cluster, the same error messages that would normally be displayed in the GUI are dumped into a text file in the current working directory on the cluster. Different HPC systems will have different conventions for the formatting of these text files, so it is important to check the specifics of your HPC system. A separate text file is created for each job run on the cluster. Thus, when initially moving from local code development to HPC analysis, it was important to run analyses on only a small subset of data in order to avoid generating an unwieldy number of error text files. In a similar fashion to the error messages, anything that would normally be displayed as text output in GUI-based interactive use of Matlab is dumped into an output text file when running code using an HPC cluster. Notice that throughout all four layers of code, frequent usage of the disp() function created outputs that were printed into the output text file. Combining the output text file and the error text file, it was possible to know how far the analysis had gotten before any error and exactly what error had caused it to stop.

Having identified the exact piece of code that caused the error, it was then possible to recreate the error on a local machine in GUI-based interactive mode. This was done by setting the jobID to the number displayed in the relevant text output file (to specify the data to be analyzed), setting the prefix for local processing, placing a debug stop point in the pipeline function prior to the point of the error, and then running the pipeline wrapper code to call the pipeline function. This procedure made it possible to examine and interact with the Matlab workspace at the time of the error, which had initially occurred on the cluster. The ability to reproduce errors that occurred during batch processing in an interactive Matlab session was critical to facilitating debugging.

Errors also occurred when jobs failed to finish or execute properly on the cluster. These errors often occurred when a job required more time or memory than had been allotted to it. These errors were not caused because of a problem with the code. Instead, the fix for these errors was often to rerun the analysis. However, for analysis steps with a very high job count, it was not efficient to rerun the entire analysis. It was also difficult if a large number of jobs had completed to know whether a small handful may not have completed properly. Thus, it was necessary to audit the analysis outputs.

The Matlab files, audit_singleChanAll.m and PACaudit.m work together to check whether phase amplitude coupling analysis has completed properly across a set of 88,160 individual EEG channel files. As explained below, for the bulk of EEG metrics analyzed in this study, subject data files were split apart into individual channel data files. This facilitated parallel processing, but it also made it impossible to check all of the output and error text files associated with running an analysis on all channel data files. The auditing code specified in these two Matlab files checked through all 88,160 files and returned error codes organized as a 1-dimensional vector across the entire set of files, making it much easier for the user to assess progress and target particular data files for reprocessing or extra scrutiny. The audit code in these files took about 3 hours to run on a local machine capable of running 8 parallel processes, but if audit_singleChanAll.m and PACaudit.m are thought of as pipeline wrapper and pipeline functions respectively, then it would be simple to modify these for implementation on an HPC cluster through the addition of a job initiation bash script. Thus, one could audit the output of an HPC analysis using another HPC analysis if one’s local machine is not powerful enough or the dataset in question is too large to run audit operations locally. Audit code was able to detect errors, but it was also useful for checking the progress of an analysis as it ran.

#### 2.3 Modularity in coding

Coding modularly facilitates analysis development, makes errors easier to find, and increases the flexibility of code. To accomplish this, code writing was easiest to implement using a similar approach to debugging. For example, when writing a new computation function to perform a power spectrum decomposition, it was easiest to work on a local machine, set a debug stop point in the pipeline function at the point that the power spectrum decomposition was to be performed, and then initiate analysis from the pipeline wrapper. Once the code stopped at the debug point, it was then easy to see exactly what variables were available in the workspace and to implement code to perform the power spectrum decomposition given those variables. The new analysis code could be written in a typical trial and error fashion interactively. Then, once the analysis was working, it could be packaged into a new power spectrum decomposition function and separated into its own file separate from the pipeline function. In this way, it was easy to remove or add analyses to the pipeline. It was also easy to focus thinking on the development of only one piece of code at a time.

#### 2.4 Frequent output saving and progress checking

When running large analyses on an HPC cluster, it was important to save interim results frequently and to check for previously completed computations. To do this, each computation function checked whether its output variables were already available before performing computation and only performed computation if the outputs were not already available. In addition, each computation function saved its outputs after completing computation (as indicated by the blue and green arrows pointing to the saved outputs from computation functions in Figure S1. In this way, work was not wasted even when the full pipeline failed to complete, a new computation function was added, or a computation function was edited. Without doing progress checks and saving outputs, it would not have been possible to complete this analysis in a reasonable amount of time.

### 3. Key steps of the present analysis

The present analysis was split into 7 functional steps: data harmonization, multi-channel analysis using HPC, splitting data into single channels, single-channel analysis using HPC, combining single channel results, combining subject results, inferential statistics. Figure S1 encompasses data harmonization, multi-channel analysis using HPC, and splitting data into single channels. Single-channel analysis using HPC followed a structure similar to that represented in Figure S1. The key differences were that rather than starting with raw data, the analysis began with the single-channel files that were output at the end of the analysis depicted in Figure S1. In addition, because all single-channel files were in a uniform format, there was no need for multiple pipeline wrapper functions for the different datasets. Below, we describe each of the analysis steps.

#### 3.1 Data harmonization

To facilitate comparison of data collected across multiple sites, it was important to harmonize data. Harmonization also helped to streamline analysis since once data were formatted in a standard way, the same analysis code could be applied to all of the data. To harmonize the data, it was necessary to write a custom job initiation and pipeline wrapper script for each dataset. As explained above (section 1), these scripts requested resources, organized metadata, specified file paths, and called the pipeline script. The main reason for writing different scripts for each dataset was to deal with the different file structures in which their raw data were stored and the different locations where their metadata could be found. We chose to include only metadata that were available across all datasets in order to facilitate comparison.

The only other aspect of analysis that was customized for each dataset was the readEEGdat.m function, which included separate code blocks for loading raw data in different formats. All data were formated into a standard EEGLAB format ^8^. readEEGdat.m was called from within the multi-channel analysis pipeline function (setReadInAggregate.m)

#### 3.2 Multi-channel HPC analysis

Although single channel analysis is more efficient for parallelization (section 4), certain analyses are fundamentally multi-channel. Thus, we implemented two rounds of HPC analysis. First, multi-channel analysis was done on data from whole subjects. Next, single-channel analysis was done on data broken down to the single channel level (sections 3.3 and 3.4).

Multi-channel analysis included the initial read in of the raw data, formatting the data, automated cleaning of the data, interpolation to a standard montage, connectivity analysis (intersite phase clustering), and splitting the data into single channels. Each of these operations was accomplished by a separate computation function within the setReadInAggregate.m pipeline function.

#### 3.3 Splitting data into single channels

The final step of the setReadInAggregate.m pipeline function was to split the data into single channel files. The key things to consider when creating single channel files were creation of unique and systematically reproducible file names and sufficient metadata to make it easy to reaggregate all single channels associated with a given subject back together after analysis. In this analysis, we chose to store single channel files alongside the subject’s raw data. Thus, the file directory of the raw data could be used to locate all of its associated channel files. This directory was stored in the summary data file associated with the subject, and each subject’s summary data file then served as the core record for all analyses associated with that subject.

#### 3.4 Single-channel HPC analysis

The bulk of EEG variables considered in the present study were computed on single channel data. These were accomplished through the construction of a single set of job initiation, pipeline wrapper, and pipeline code files to handle analysis for all channels. By using the same set of code for all channels regardless of which dataset they had originally come from, it was possible to be certain of uniformity in analysis methods.

Within the single channel pipeline, there were several computation functions. Specifically, we calculated power spectra, 1/f slope and intercept, alpha peak frequency, phase amplitude coupling, and multi-scale sample entropy.

#### 3.5 Combining single channel results

Because the naming convention and save location of single channel files was determined by the information contained in the subject’s summary data file, it was possible to load the summary data file and use its metadata to loop across the subject’s associated single channel files. This allowed the subject level summary files to be updated with the results of the single channel analysis. Example code for performing this combination step can be seen in stitchFilesTogether.m.

#### 3.6 Combining subject results

Considering the large number of variables extracted from each subject’s data, we next sought to create standardized output variables that binned data across frequency and head topography. This was accomplished through the use of the extractingFinalVariables.m script. This script read in each subject’s summary output data and then pulled a standard set of output variables. This step allowed data from all subjects to be represented in a single .csv file to facilitate inferential statistics.

Notice in both this and the previous section, the code used to implement the analysis step does not perform any actual computation or interact with the raw data in any way. As a result, these steps require much less computation time than the main analysis steps. Using the file path conventions discussed above, it was easy to write code for these steps that could run either locally or on the HPC cluster. However, in practice, these steps could be run locally in a reasonable amount of time.

#### 3.7 Inferential statistics

Finally, the .csv file generated in the combining subject results step was imported into R for final inferential statistical analysis. At this point, the .csv file could be imported into any environment for statistical analysis depending on the user’s preference. Our inferential statistics were carried out using the script effectSizeCalculations.R.

### 4. Why splitting to single channels is so helpful on an HPC cluster

It is common practice to analyze data using a process that amounts to two nested loops. One loop is done across subjects. The second loop is done across data channels. This style of analysis does not take full advantage of the benefits of HPC analysis because of its poor compatibility with parallelization and its heavy memory usage. Take incompatibility with parallelization first, the power of HPC analysis comes from the ability to use many computing nodes simultaneously. If only one node is being asked to loop through all of the data, then there will be no speed gained from HPC use. For example, in the present analysis, we found that because of the bootstrapping required for calculating phase amplitude coupling, a single channel required approximately 1 hour of computing time. Thus, for a subject with 32 channels of data, analysis would take 1.5 days. For a typical study in which analysis is done as data are collected, this processing time may be acceptable. However, for the present study with 776 participants, this meant that approximately 3 years of compute time would have been needed to loop across all subjects and channels one at a time. This example suggests that simply parallelizing over subjects may solve the problem. If it were possible to process all subjects at the same time, then the entire analysis would only take 1.5 days to run, no matter how many subjects were in the study. However, this will run into problems with memory usage.

Typical desktop computers used for data analysis have 32 GB of memory, and in many cases they may have as much as 128 GB. Thus, when performing only one process at a time on a local machine, there is generally little need to worry about memory usage. However, when working with HPC clusters, there is often only 3-5 GB of memory per processor. The ShARC cluster used in the present study has 4 GB of memory per processor. This means that analyses which require more than 4 GB of memory will take memory resources away from other processors and be difficult for the HPC scheduling software to carry out while balancing demands from other users. In the case of using the ShARC, we found that when requesting less than 4 GB of memory per analysis job, it was possible to receive allocation to as many as 1000 simultaneous computer processors. By contrast, when requesting 64 GB of memory, it was rare to receive more than 4 computer processors. Thus, memory limitations are a primary reason why it is important to break out of both the subject and the channel loop when structuring analysis.

## EEG variables best predicted by age, sex, and diagnosis

For each age group and predictor, the best predicted EEG variable was plotted in Supplemental Figure 2 for visual inspection of the prediction variables’ efficacy. IQ’s best predicted EEG measures are not shown as IQ was a weak predictor and, unlike ASD diagnosis, not theoretically motivated in this work. For age, the average MSE across all scales in a central scalp region was the best predicted variable in the youngest age group (Supplemental Figure 2a); the log of theta power in a left lateral region was the best predicted variable in the middle age group (Supplemental Figure 2b), and the log of delta power in the right hemisphere was the best predicted variable in the oldest age group (Supplemental Figure 2c). For sex, short range theta band connectivity was the best predicted variable in the youngest age group (Supplemental Figure 2d); the difference between left medial and left lateral low gamma power was the best predicted variable in the middle age group (Supplemental Figure 2e), and the log of delta power in the left lateral region was the best predicted variable in the oldest age group (Supplemental Figure 2f). Finally, for diagnosis, the difference between medial and lateral measurements of normalized PAC between the alpha and high gamma bands was the best predicted variable in the youngest age group (Supplemental Figure 2g); delta power in a left lateral region was the best predicted variable in the middle age group (Supplemental Figure 2h), and the difference in alpha power between right medial and right lateral channels was the best predicted variable in the oldest age group (Supplemental Figure 2i).

---

**Supplemental Figure 1.**
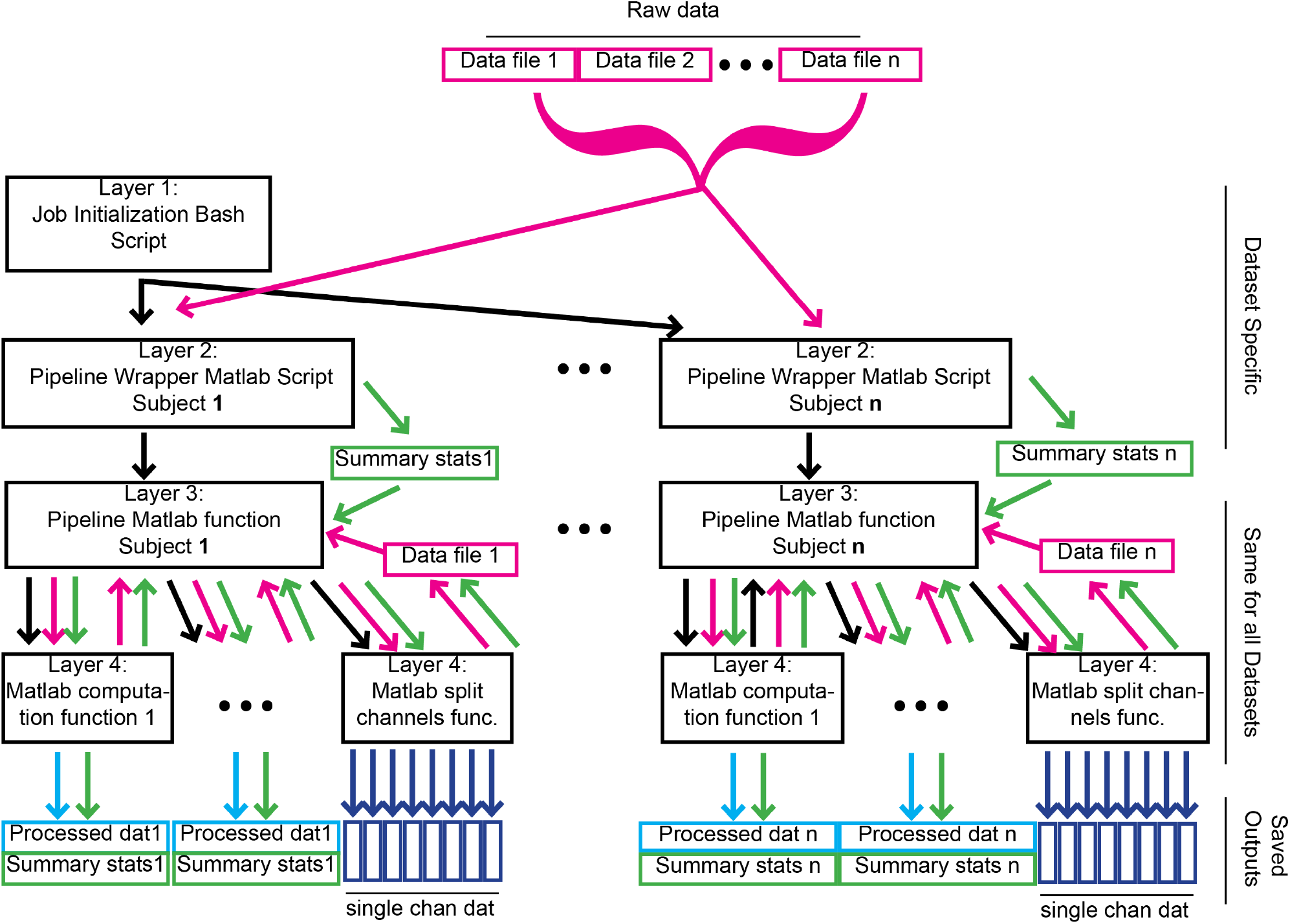
General outline of multichannel analysis using HPC cluster. Code was organized into four layers. The top layer, the job initialization bash script, was called from the HPC linux command line. It requested resources from the cluster, specified the number of data files to be processed, initiated one job on the cluster per data file, opened Matlab, and called the pipeline wrapper script with the jobID number as an input. The pipeline wrapper Matlab script handled file path control, organized metadata, and called the pipeline function. The pipeline function loaded the raw data into Matlab and then submitted the raw data to a series of processing steps. Each processing step was executed by a different Matlab computation function. These computation functions were modular and could be easily plugged in or removed from the analysis pipeline. Black arrows indicate the flow of code based calls from one script or function to another. Green lines indicate the flow of subject specific summary statistics files. Pink lines indicate the flow of raw data. Light blue lines indicate the flow of processed data. Dark blue lines indicate the flow of single channel data. Ellipses within the fourth layer of code indicate that an arbitrary number of additional computation functions could be inserted within the pipeline. Ellipses between the left and right sides of the flow chart indicate that an arbitrary number of separate subject files could be processed in parallel.

**Supplemental Figure 2.**
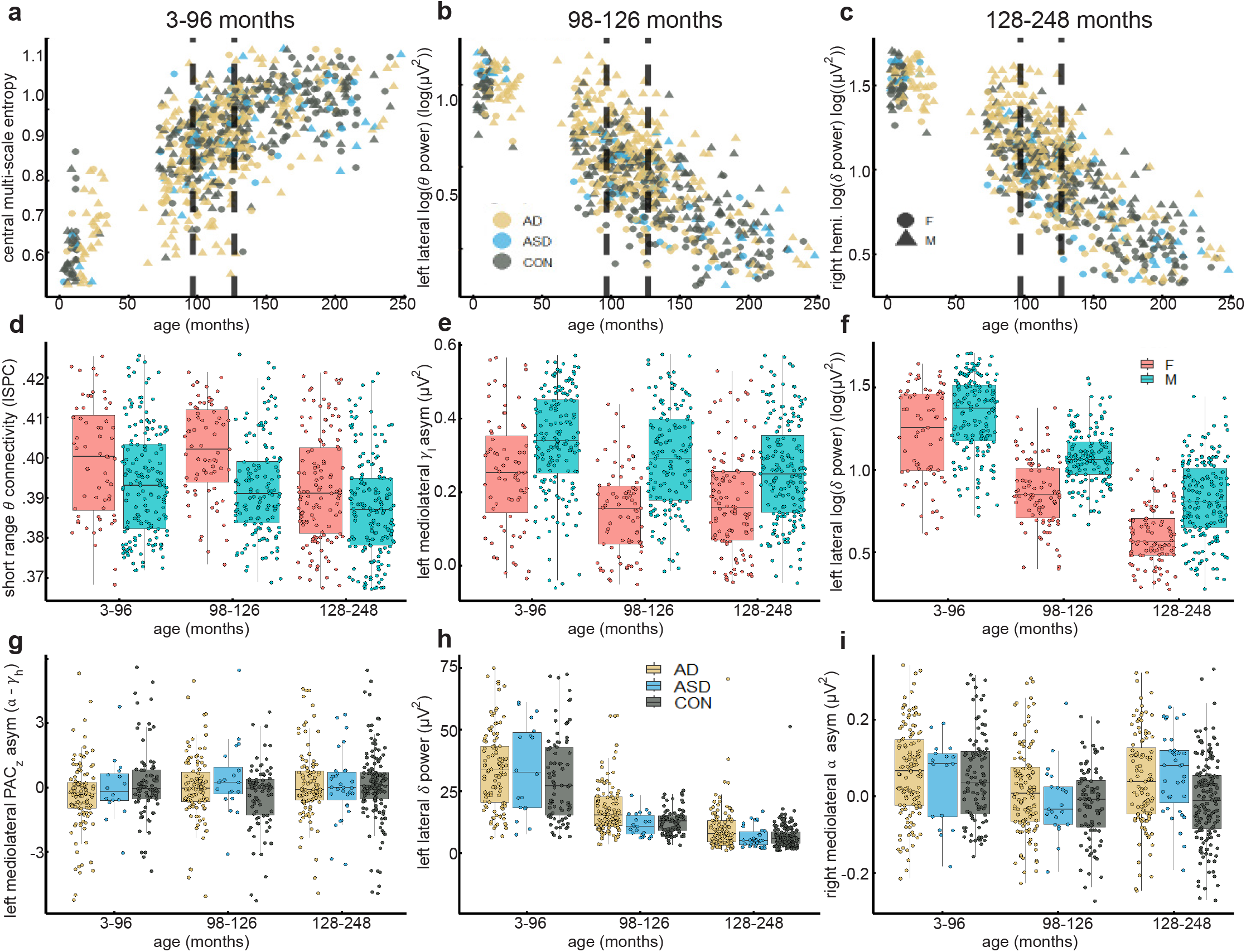
The best predicted EEG measures are displayed for each predictor within each age group. All data from both training and test sets were pooled for these plots. **a-c.** Scatter plots display age on the x-axis and different dependent variables on the y-axis. See y-axis labels for dependent variables and units. Dependent variables are the variables that were best predicted by age in each of the three age groups respectively. Vertical dashed lines indicate cut offs between age groups. Colors indicate diagnosis group and shapes indicate sex (see keys). **d-f.** Box plots display age group on the x-axis and different dependent variables on the y-axis. See y-axis labels for dependent variables and units. Dependent variables are the variables that were best predicted by sex in each of the three age groups respectively. Colors indicate sex (see key). For both age and sex as predictors, although each dependent variable was chosen based on independent analysis in only one age group, visual inspection reveals that differences as a function of age and sex were present for these variables across all age groups. **g-i.** Box plots are formatted similarly to **d-f** except the dependent variables displayed on the y-axes were those best predicted by autism diagnosis. Colors indicate autism diagnosis (see key). Note that unlike age and sex, when diagnosis is used as a predictor, effects appear limited to the age group in which they were detected. For example, in panel **g** autistic participants exhibit lower mediolateral asymmetry of z-scored phase-amplitude coupling between the alpha and low gamma bands than controls in the youngest age group, but this pattern is not present in the two older age groups. Similar age-selective patterns are evident in panels **h** and **i** for the dependent variables best predicted by diagnosis in the older age groups.

**Table S1:**
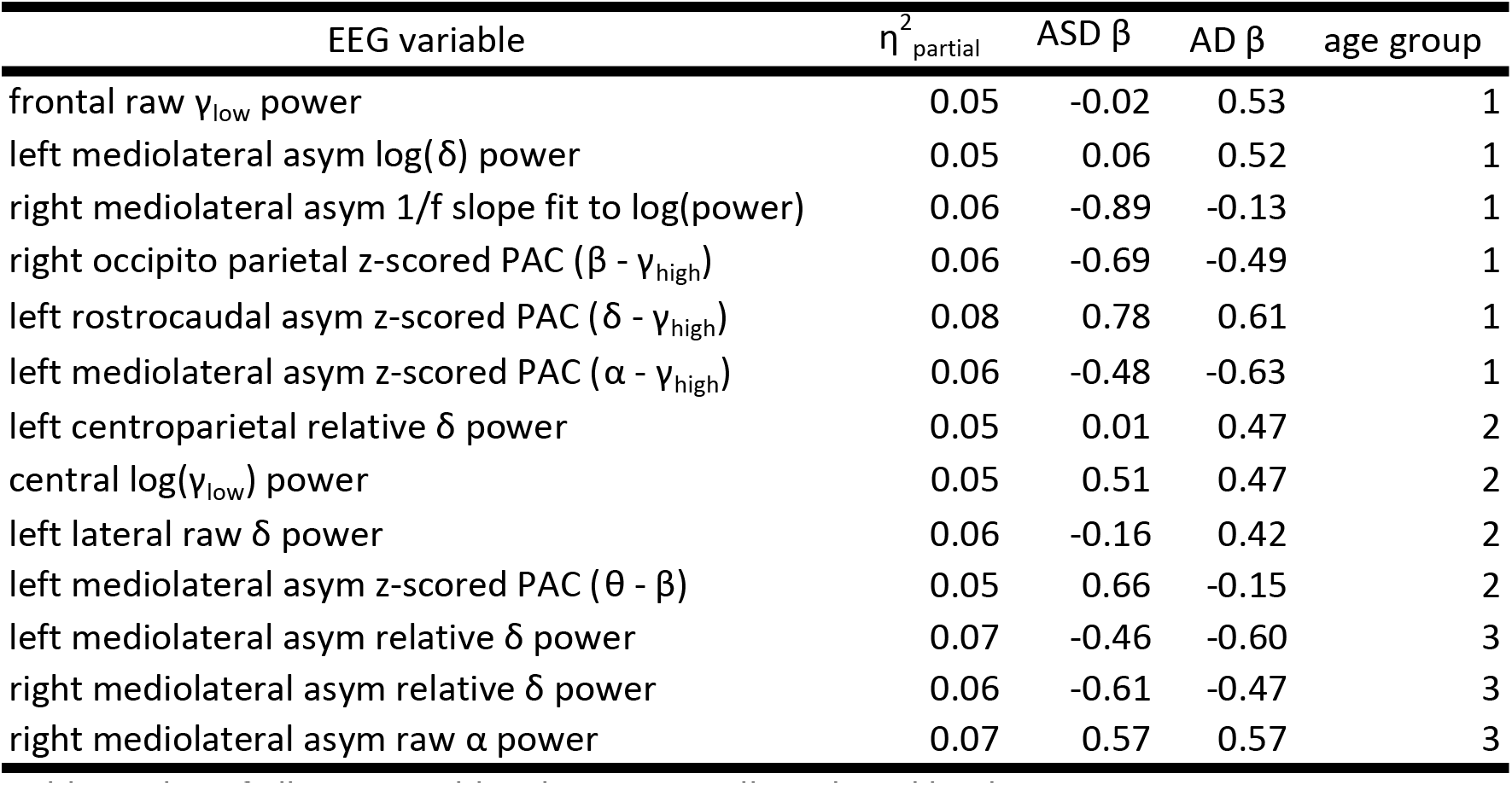
list of all EEG variables that were well predicted by diagnosis.

| EEG variable | $\eta^2_{\text{partial}}$ | ASD $\beta$ | AD $\beta$ | age group |
| --- | --- | --- | --- | --- |
| frontal raw $\gamma_{\text{low}}$ power | 0.05 | -0.02 | 0.53 | 1 |
| left mediolateral asym log($\delta$) power | 0.05 | 0.06 | 0.52 | 1 |
| right mediolateral asym 1/f slope fit to log(power) | 0.06 | -0.89 | -0.13 | 1 |
| right occipito parietal z-scored PAC ($\beta$ - $\gamma_{\text{high}}$) | 0.06 | -0.69 | -0.49 | 1 |
| left rostrocaudal asym z-scored PAC ($\delta$ - $\gamma_{\text{high}}$) | 0.08 | 0.78 | 0.61 | 1 |
| left mediolateral asym z-scored PAC ($\alpha$ - $\gamma_{\text{high}}$) | 0.06 | -0.48 | -0.63 | 1 |
| left centroparietal relative $\delta$ power | 0.05 | 0.01 | 0.47 | 2 |
| central log($\gamma_{\text{low}}$) power | 0.05 | 0.51 | 0.47 | 2 |
| left lateral raw $\delta$ power | 0.06 | -0.16 | 0.42 | 2 |
| left mediolateral asym z-scored PAC ($\theta$ - $\beta$) | 0.05 | 0.66 | -0.15 | 2 |
| left mediolateral asym relative $\delta$ power | 0.07 | -0.46 | -0.60 | 3 |
| right mediolateral asym relative $\delta$ power | 0.06 | -0.61 | -0.47 | 3 |
| right mediolateral asym raw $\alpha$ power | 0.07 | 0.57 | 0.57 | 3 |

## Notes

### Competing Interest Statement

The authors have declared no competing interest.

### Funding Statement

This study did not receive any funding

### Author Declarations

National Institute of Mental Health Data Archive (NDA) data repository 10.15154/1528473 was used for this study. This repository was created by the first author using the NDA data selection tool.

